# Development and validation of a machine learning model for predicting illness trajectory and hospital resource utilization of COVID-19 hospitalized patients – a nationwide study

**DOI:** 10.1101/2020.09.04.20185645

**Authors:** Michael Roimi, Rom Gutman, Jonathan Somer, Asaf Ben Arie, Ido Calman, Yaron Bar-Lavie, Udi Gelbshtein, Sigal Liverant-Taub, Arnona Ziv, Danny Eytan, Malka Gorfine, Uri Shalit

## Abstract

**Background:** The spread of COVID-19 has led to a severe strain on hospital capacity in many countries. There is a need for a model to help planners assess expected COVID-19 hospital resource utilization.

**Methods:** Retrospective nationwide cohort study following the day-by-day clinical status of all hospitalized COVID-19 patients in Israel from March 1st to May 2nd, 2020. Patient clinical course was modelled with a machine learning approach based on a set of multistate Cox regression-based models with adjustments for right censoring, recurrent events, competing events, left truncation, and time-dependent covariates. The model predicts the patient’s entire disease course in terms of clinical states, from which we derive the patient’s hospital length-of-stay, length-of-stay in critical state, the risk of in-hospital mortality, and total and critical care hospital-bed utilization. Accuracy assessed over eight cross-validation cohorts of size 330, using per-day Mean Absolute Error (MAE) of predicted hospital utilization averaged over 64 days; and area under the receiver operating characteristics (AUROC) for individual risk of critical illness and in-hospital mortality, assessed on the first day of hospitalization. We present predicted hospital utilization under hypothetical incoming patient scenarios.

**Findings:** During the study period, 2,703 confirmed COVID-19 patients were hospitalized in Israel. The per-day MAEs for total and critical-care hospital-bed utilization, were 4·72 ± 1·07 and 1·68 ± 0·40 respectively; the AUROCs for prediction of the probabilities of critical illness and in-hospital mortality were 0·88 ± 0·04 and 0·96 ± 0·04, respectively. We further present the impact of several scenarios of patient influx on healthcare system utilization, and provide an R software package for predicting hospital-bed utilization.

**Interpretation:** We developed a model that, given basic easily obtained data as input, accurately predicts total and critical care hospital utilization. The model enables evaluating the impact of various patient influx scenarios on hospital utilization and planning ahead of hospital resource allocation.

**Funding:** The work was funded by the Israeli Ministry of Health. M.G. received support from the U.S.-Israel Binational Science Foundation (BSF, 2016126).

Research in context
Evidence before this study
COVID19 outbreaks are known to lead to severe case load in hospital systems, stretching resources, partially due to the long hospitalizations needed for some of the patients. There is a crucial need for tools helping planners assess future hospitalization load, taking into account the specific characteristics and heterogeneity of currently hospitalized COVID19 patients, as well as the characteristics of incoming patients. We searched PubMed for articles published up to September 9, 2020, containing the words “COVID19” and combinations of “hospital”, “utilization”, “resource”, “capacity” and “predict”. We found 145 studies; out of them, several included models that predict the future trend of hospitalizations using compartment models (e.g. SIR models), or by using exponential or logistic models. We discuss two of the more prominent ones, which model explicitly the passage of patients through the ICU. These models (i) do not take into account individual patient characteristics; (ii) do not consider length-of-stay heterogeneity, despite the fact that bed utilization is in part determined by a long tail of patients requiring significantly longer stays than others; (iii) do not correct for competing risks bias. We further searched for studies containing the words “COVID19” and “multistate”, and “COVID19” and “length” and “stay”. Out of 317 papers, we found two using multistate models focusing only on patients undergoing ECMO treatment.

Added value of this study
We present the first model predicting hospital load based on the individual characteristics of hospitalized patients: age, sex, clinical state, and time already spent in-hospital. We combine this with scenarios for incoming patients, allowing for variations by age, sex and clinical state. The model’s precise predictions are based on a large sample of complete, day-by-day disease trajectories of patients, with a full coverage of the entire COVID-19 hospitalized population in Israel up to early May, 2020 (*n* =2, 703). We provide the model, as well as software for fitting such a model to local data, and an anonymized version of the dataset used to create the model.

Implications of all the available evidence
Accurate predictions for hospital utilization can be made based on easy to obtain patient data: age, sex, and patient clinical state (moderate, severe or critical). The model allows hospital-and regional-level planners to allocate resources in a timely manner, preparing for different patient influx scenarios.

## 1. Introduction

The coronavirus disease 2019 (COVID-19) was first recognized in Wuhan, China in December 2019. On March 11 2020 the world health organization characterized the disease as a pandemic^1^ Worldwide, the COVID-19 pandemic poses a major challenge for the healthcare systems and it will probably continue to pose challenges in the coming years^2,3^ In particular, the disease has taken its toll on healthcare systems around the world with some patients requiring lengthy general and intensive care^4^.

Given the danger of unprecedented burden on healthcare systems due to COVID-19, there is a need for tools that help decision-makers plan resource allocation on the unit, hospital, regional and national levels. In this study we aimed to use a nationwide hospitalization registry which includes the day-by-day hospitalization record of all the confirmed COVID-19 patients in Israel. We used the registry to develop a tool for accurate projections of the total number of hospitalized patients and critical-care occupancy based on the state of the current hospitalized patient population and projections of hospital patient influx. To facilitate use of the model, we provide an R^5^ software package^a^ enabling anyone with access to similar data to develop a model tailored to specific patient and healthcare system characteristics and provide a web-application^b^ that receives as input the characteristics of a patient and predicts the probabilities of different disease courses, including length-of-stay (LOS), probability of becoming critically ill, expected length-of-stay in critical state (LOSCS), and the probability of in-hospital mortality. Finally, we will share an anonymized version of the dataset used to develop the tool, where we shift hospitalization dates and aggregate age into 5-year categories.

## 2. Methods

We conducted a retrospective cohort study based on the Israeli Ministry of Health (MOH) COVID-19 hospitalized patient registry. The registry includes the patients’ age and sex, dates and results of their severe acute respiratory syndrome corona virus-2 (SARS-COV-2) polymerase chain reaction (PCR) tests, dates of hospital admissions and discharge, *daily* clinical status during the admission (moderate, severe or critical, as detailed below), and the death registry. We included in the analyses all the patients who were admitted between the 1st of March and the 2nd of May 2020 and who were hospitalized for at least one day. We excluded patients who were missing age or sex documentation. No data imputation was performed.

### 2.1. Outcomes

The primary outcome was prediction of total and critical-care bed occupancy on a calendar scale, where occupancy is due to currently hospitalized patients staying in the hospital, as well as due to newly arriving patients. We do this by predicting for each patient their day-by-day clinical state, including days in which the patient is in a critical state, non-critical, discharged, or possibly died. We further use the day-by-day clinical state predictions for predicting the overall risk for a single patient of being in a critical state at some point throughout hospitalization, risk of in-hospital mortality, expected hospital LOS and expected LOSCS.

### 2.2. Definitions

COVID-19 confirmed diagnosis was defined as a patient who was found to be positive for SARS-CoV-2 PCR tests^6^ Patients’ clinical state during admission (mild or moderate, severe, and critical) was defined by the Israeli MOH guidelines, which are closely related to the National Institute for Health treatment guidelines^7^.A *mild* or *moderate* clinical state was defined as patients with symptoms such as fever, cough, sore throat, malaise, headache, or muscle pain, without shortness of breath, dyspnea on exertion, or abnormal imaging, or as patients with clinical or imaging evidence for lower respiratory disease and the saturation of oxygen (SpO_2_) > 90% on room air. For brevity we will refer to this state as *moderate* henceforth. A *severe* clinical state was defined as a patient with a respiratory rate higher than 30 breaths per minute, SpO_2_ < 90% on room air, or ratio of the arterial partial pressure of oxygen to fraction of inspired oxygen (PaO_2_/FiO_2_) < 300 mmHg. A *critical* clinical state was defined as a state where the patient suffers from respiratory failure which requires invasive/non-invasive mechanical ventilation, septic shock, or multiorgan dysfunction. In addition, we denote patients who were discharged from the hospital to their home or to out-of-hospital quarantine as *Discharged*. We note that discharged patients might be readmitted upon deterioration.

### 2.3. Statistical analysis methods

The clinical state of COVID-19 patients often alternates between moderate and more severe clinical states; see table S1 in supplementary materials (SM) for descriptive statistics of these transitions between clinical states. We modelled the way patients move between the different clinical states over time using a statistical machine learning approach. Specifically, we used an approach whose purpose is exactly to model such processes: a multistate Cox regression-based survival analysis with right censoring, competing events, recurrent events, left truncation, and time-dependent covariates, to model the way patients move between the different clinical states over time^8–11^ The multistate model has four states: (1) moderate or severe, (2) critical, (3) discharged and (4) deceased. We chose to merge the moderate and severe clinical states due to sample size considerations. The model consists of six semiparametric Cox regression models, one for each possible state-to-state transition, as depicted in Figure 1; some transitions were excluded as either clinically implausible or due to few observed transitions. For details see SM sections S1.1 – S1.3.

**Figure 1:**
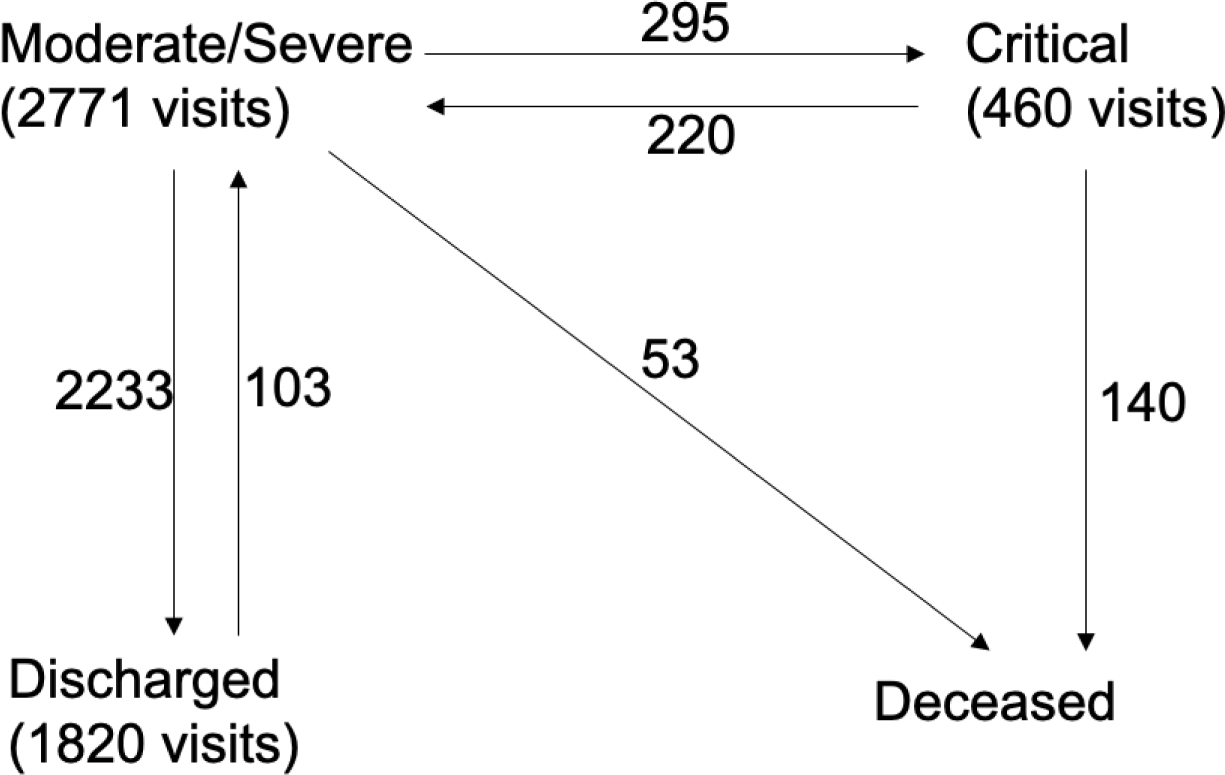
Multistate model. We model a COVID-19 patient’s disease course as moving between four possible states: (i) moderate or severe, (ii) critical, (iii) discharged and (iv) deceased. We combined the two clinical states moderate and severe into a single model state due to statistical considerations; however, we emphasize that we keep a distinction between the two by a covariate indicating whether the patient first entered at mild/moderate clinical state or at a severe clinical state. Numbers next to arrows indicate number of observed transitions; each patient can make several state transitions, and may visit a transient state more than once.

The six semiparametric models each took in age, sex, and state at hospitalization as covariates; for the latter we kept the distinction between moderate and severe clinical states. We also added time-dependent covariates encoding the hospitalization history of the patient: cumulative days in hospital, and whether the patient had been in a critical state before; see SM section S1.1.

Since hospitalization consists of potentially multiple transitions between transient states, the absolute risks, also known as the cumulative incidence functions, do not have a tractable analytic form. Thus, we employed a method called Monte-Carlo (MC) sampling for obtaining unbiased estimates of patient and cohort statistics from the multistate model. Each MC sample for a given patient consists of a disease course, in terms of clinical states over time and how much time is spent in each one, conditioned on the patient’s history and covariates. We calculated statistics such as median LOS or expected number of hospitalized patients per day based on aggregating the results of 20,000 MC samples for each patient. Standard errors were obtained by weighted bootstrap. See SM sections S1.4 and S1.7.

### 2.4. Model validation – hospital resource utilization and individual patient disease course prediction

We validated our model using 8-fold cross-validation – fitting the model on 7/8 of the data and evaluating performance on the remaining held-out 1/8, repeated eight times. Each held-out set consisted of 330 patients.

We validated predictions of hospital utilization by two methods: “Snapshot”, where we define a start date (we chose either April 1st or April 15th, 2020), and predict the future resource utilization for the set of all patients who are in-hospital on that date, without taking into account future incoming patients; and “Arrival process”, where we use the known hospitalization dates and characteristics of incoming patients between March 1st to May 2nd 2020 in order to estimate utilization for the entire course of the “first wave” in Israel; see SM section S1.3 for description of both. For both validation methods we estimate the Mean Absolute Error (MAE) between the model’s predictions per-day and the actual number of hospitalized (or critical) patients on that day; the mean is over the number of days in the prediction window; see SM section S1.3.

We further validate the model’s performance by testing its predictions on the level of the individual patient: we use data from the first day of a patient’s admission to predict their probability of becoming critically ill (among patients who were non-critically ill on their 1st day) and the probability of in-hospital mortality. For both outcomes we report Area Under Receiver Operating Characteristic (AUROC) with inverse weighting correction for censoring, see SM section S1.3 for details. Finally, we validated the calibration of our predictions by tracking expected number of deaths vs. actual number of deaths over time in an “Arrival+Snapshot” scenario, see SM section S1.3 for details.

### 2.5. Using the model for prediction of hospital utilization under hypothetical scenarios

In order to illustrate how our model may be employed for utilization prediction, we focus on a single held-out cohort of 330 patients. For this cohort we predict the total future hospital-bed and critical hospital-bed utilization up to 49 days ahead, starting from March 15th. Utilization for this cohort is composed of patients among the 330 who were hospitalized at the starting date and remain at the hospital, as well as utilization by newly arrived patients.

We present the expected hospital utilization and the number of deaths under three putative patient arrival scenarios (i) “younger”: rate and state of incoming patients are the same as in Israel during the weeks from March 15th to May 2nd, but all patients ages 60+ are replaced with patients in their 40s and 50s; (ii) “milder”: rate and age of incoming patients are the same as in Israel during the weeks from March 15th to May 2nd, but all patients incoming only in moderate or severe clinical state, none at critical, and (iii) “eldercare nursing home (NH) outbreak” where we assume that in addition to the arrival of patients as happened in Israel from March 15th to May 2nd there is a single week during which there are four times as many incoming patients aged 70+, arriving in various clinical states. The details of the scenarios are given in SM section S2.4.

## 3. Results

### 3.1. Hospitalized patient characteristics

The first patient with COVID-19 In Israel was diagnosed on February 27th 2020. As of May 2nd 2020, 16,137 patients had confirmed positive diagnosis. The median (IQR) age of the confirmed patients was 33 (21-54); 44·5% were females. Out of the 16,137 confirmed COVID-19 patients up to May 2nd, 2,703 (17%) were hospitalized by May 2nd for at least one full day^c^ Of these 2,703 hospitalized patients, 28 patients had no documented age or sex covariates. For the remaining cohort of 2,675 patients median (IQR) age was 58 (39-73), 44·19% were female. The demographics, medical history, and the clinical status of hospitalized patients are shown in Table 1. We use the model to estimate the median, 10% and 90% quantiles of the length-of-stay for hospitalized patients stratified by clinical state at time of admission. The results are shown in Figure 2 and Table S7 in the SM; in Table S8 in SM we report LOSCS results.

**Figure 2:**
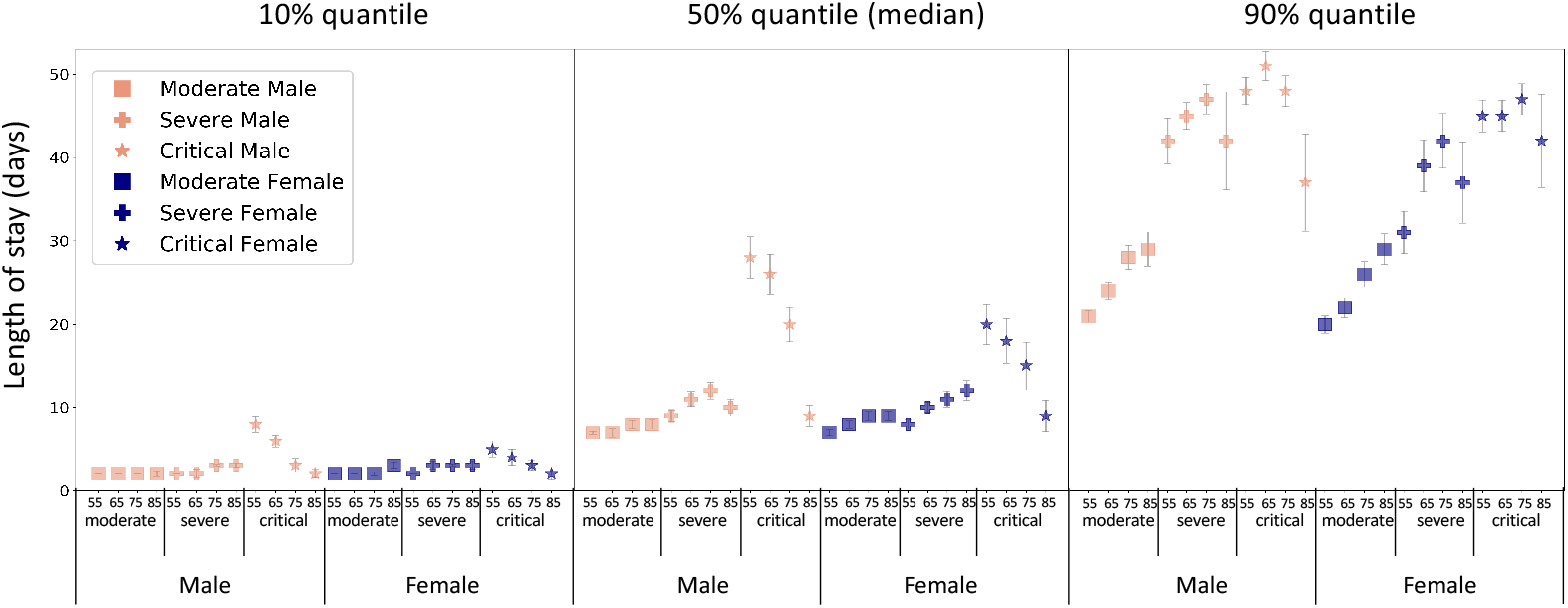
LOS stratified by patients’ age, sex and the clinical state at the time of admission. Model-based estimates of quantiles of length of stay in days based on 20,000 Monte Carlo results for each patient strata. Error bars indicate 95% confidence interval, calculated by weighted bootstrap.

**Table 1:**
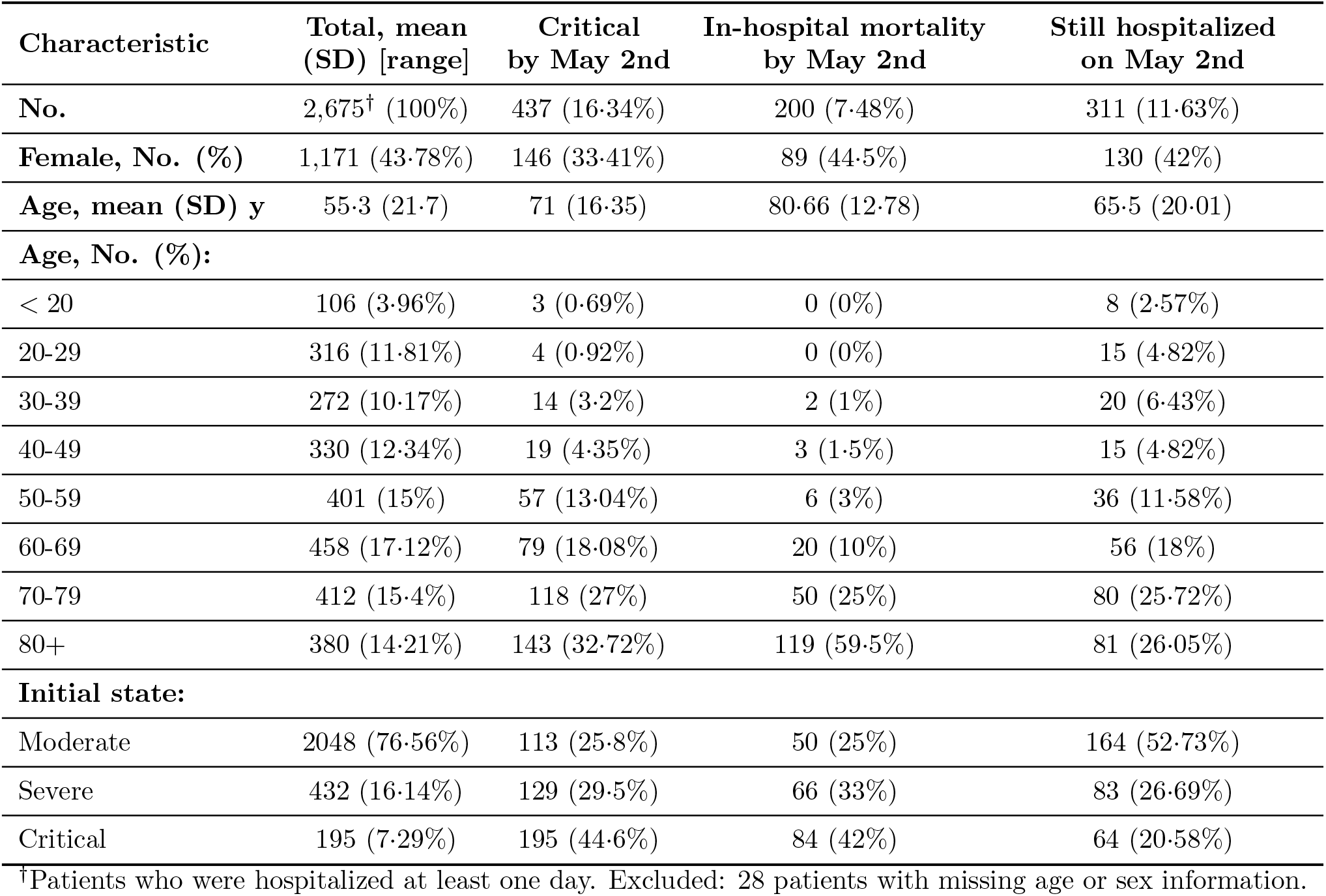
Demographics and clinical characteristics of patients in the Israeli COVID-19 registry who were Hospitalized between March 1st and May 2nd.

| Characteristic | Total, mean (SD) [range] | Critical by May 2nd | In-hospital mortality by May 2nd | Still hospitalized on May 2nd |
| --- | --- | --- | --- | --- |
| **No.** | 2,675<sup>†</sup> (100%) | 437 (16.34%) | 200 (7.48%) | 311 (11.63%) |
| **Female, No. (%)** | 1,171 (43.78%) | 146 (33.41%) | 89 (44.5%) | 130 (42%) |
| **Age, mean (SD) y** | 55.3 (21.7) | 71 (16.35) | 80.66 (12.78) | 65.5 (20.01) |
| **Age, No. (%):** |  |  |  |  |
| < 20 | 106 (3.96%) | 3 (0.69%) | 0 (0%) | 8 (2.57%) |
| 20-29 | 316 (11.81%) | 4 (0.92%) | 0 (0%) | 15 (4.82%) |
| 30-39 | 272 (10.17%) | 14 (3.2%) | 2 (1%) | 20 (6.43%) |
| 40-49 | 330 (12.34%) | 19 (4.35%) | 3 (1.5%) | 15 (4.82%) |
| 50-59 | 401 (15%) | 57 (13.04%) | 6 (3%) | 36 (11.58%) |
| 60-69 | 458 (17.12%) | 79 (18.08%) | 20 (10%) | 56 (18%) |
| 70-79 | 412 (15.4%) | 118 (27%) | 50 (25%) | 80 (25.72%) |
| 80+ | 380 (14.21%) | 143 (32.72%) | 119 (59.5%) | 81 (26.05%) |
| **Initial state:** |  |  |  |  |
| Moderate | 2048 (76.56%) | 113 (25.8%) | 50 (25%) | 164 (52.73%) |
| Severe | 432 (16.14%) | 129 (29.5%) | 66 (33%) | 83 (26.69%) |
| Critical | 195 (7.29%) | 195 (44.6%) | 84 (42%) | 64 (20.58%) |

<sup>†</sup>Patients who were hospitalized at least one day. Excluded: 28 patients with missing age or sex information.

In Table 2 we present the probability of different patient populations entering a critical state and the probability for in-hospital mortality, as estimated by the model. Both probabilities sharply increase with age; males of all ages tend to have a greater probability of becoming critically ill compared to females entering the hospital at the same clinical state, but hospitalized Females over age 75 tend to have higher risk of mortality compared to hospitalized males entering at comparable age and clinical state. Probabilities and LOS and LOSCS quantiles for younger ages are given in Tables S5-S8 in the SM; for cumulative distribution function of LOS see Figure S2 in the SM.

**Table 2:** Probability of death and probability of becoming critical stratified by age, sex and clinical state.

| *Incoming state, age* | Probability of in-hospital mortality |  | Probability of becoming critical |  |
| --- | --- | --- | --- | --- |
|  | *Male* | *Female* | *Male* | *Female* |
| Moderate, 55 | 0.9%<sup>‡</sup><br>(0.2%, 1.5%) | 1.3%<br>(0.4%, 2.3%) | 5.9%<br>(4.6%, 7.3%) | 4.1%<br>(2.5%, 5.7%) |
| Moderate, 65 | 2.2%<br>(1.2%, 3.2%) | 2.6%<br>(1.2%, 4%) | 8.5%<br>(6.9%, 10.1%) | 6.1%<br>(4.1%, 8%) |
| Moderate, 75 | 5.8%<br>(3.4%, 7.8%) | 5.3%<br>(3%, 7.6%) | 12.9%<br>(10.5%, 15.2%) | 9.9%<br>(6.9%, 12.8%) |
| Moderate, 85 | 15.1%<br>(8.9%, 21.2%) | 12.2%<br>(7.6%, 16.7%) | 17.9%<br>(13.9%, 21.9%) | 13.4%<br>(8.9%, 17.9%) |
| Severe, 55 | 4.3%<br>(0%, 9.5%) | 7.5%<br>(1.2%, 13.7%) | 24.1%<br>(18.7%, 29.4%) | 18.5%<br>(12.8%, 24.2%) |
| Severe, 65 | 9.6%<br>(3.9%, 15.2%) | 12.6%<br>(4.3%, 20.8%) | 31.9%<br>(26.7%, 37.0%) | 25.6%<br>(20.1%, 31.1%) |
| Severe, 75 | 21.4%<br>(14.9%, 28%) | 21.7%<br>(12.6%, 30.9%) | 40.4%<br>(35%, 45.7%) | 32.3%<br>(26.7%, 37.9%) |
| Severe, 85 | 44.3%<br>(32.3%, 56.3%) | 38.9%<br>(28.8%, 48.9%) | 46.7%<br>(39.3%, 56.3%) | 38.5%<br>(31.1%, 45.8%) |
| Critical, 55 | 15.3%<br>(3.5%, 27.2%) | 29.9%<br>(10.8%, 49%) | 100% | 100% |
| Critical, 65 | 29.9%<br>(17.9%, 41.9%) | 41.4%<br>(25.5%, 57.2%) | 100% | 100% |
| Critical, 75 | 54.6%<br>(42.9%, 66.3%) | 54.9%<br>(42.6%, 67.1%) | 100% | 100% |
| Critical, 85 | 82.9%<br>(74.4%, 91.3%) | 75.4%<br>(65.2%, 85.7%) | 100% | 100% |

<sup>‡</sup>Probabilities are based on Monte Carlo results, with weighted bootstrap 95% confidence interval.

### 3.2. Cox models

The results for all six Cox models are given in Tables S2-S4 in the SM.

### 3.3. Model validation

The following results are all averaged over the eight held-out validation cohorts. Using “Snapshot” evaluation with April 1st as start date, MAE for predicting the per-day number of hospitalized patients is 3·15 ±1·20 for total hospital-bed utilization and 1·47±0·56 for critical-care bed utilization. Using “Snapshot” evaluation with April 15th as start data, MAE for predicting the per-day number of hospitalized patients is 3·13 ± 1·07 for total hospital-bed utilization and 1·98 ± 0·93 for critical hospital-bed utilization. Using “Arrival” evaluation, MAE for predicting the per-day number of hospitalized patients is 4·72 ± 1·07 for total hospital-bed utilization and 1·68 ± 0·40 for critical hospital-bed utilization. See Table S9 and Figures S3-S4 in the SM for full results.

Using only information from the first day of a patient’s hospitalization, the AUROC for predicting in-hospital mortality and for predicting the outcome of becoming critically ill (among patients who were non-critically ill on their first day of admission) were 0·96 ± 0·04 and 0·88 ± 0·04 respectively; see Table S10 in the SM for full results. Table 3 presents the number of deaths predicted by our model under the true patient influx process (“Expected” column), which matches very closely the observed number of deaths, showing the model is well-calibrated.

**Table 3:** Number of deaths (in-hospital mortality) within a random subset of 330 validation set (held-out) patients. “Observed” is true number of deaths, “Expected” is prediction by the model. “Younger”, “Milder” and “NH Outbreak” are hypothetical scenarios. (i) “Younger”: rate and state of incoming patients are the same as in Israel during the weeks from March 15th to May 2nd, but with patients in their 50s and 60s instead of 60+; (ii) “Milder”: rate and age of incoming patients are the same as in Israel during the weeks from March 15th to May 2nd, but all patients incoming only in moderate and severe state, none at critical, and (iii) “Nursing home (NH) outbreak” where we assume that in addition to the arrival of patients as happened in Israel from March 15th to May 2nd there is a single week during which there are four times as many incoming patients aged 70+, arriving in various clinical states.

| Days | Observed | Expected | *Hypothetical scenarios* |  |  |
| --- | --- | --- | --- | --- | --- |
|  |  |  | Younger | Milder | NH<br>Outbreak |
| 5 | 7 | 6.5 | 0.6 | 3.7 | 8.2 |
| 10 | 16 | 16.6 | 1.9 | 11.7 | 22.6 |
| 15 | 20 | 20.4 | 2.5 | 15 | 28.3 |
| 20 | 23 | 22.8 | 3 | 17.3 | 32.1 |
| 25 | 24 | 25.4 | 3.4 | 19.7 | 36.5 |
| 30 | 25 | 25.9 | 3.5 | 20.1 | 37 |
| 35 | 26 | 26.6 | 3.7 | 20.6 | 37.7 |

### 3.4. Predicting hospital-bed utilization

In Figure 3 and Table 3 we show an example of the utilization and mortality projections generated by the model for several hypothetical scenarios.

**Figure 3:**
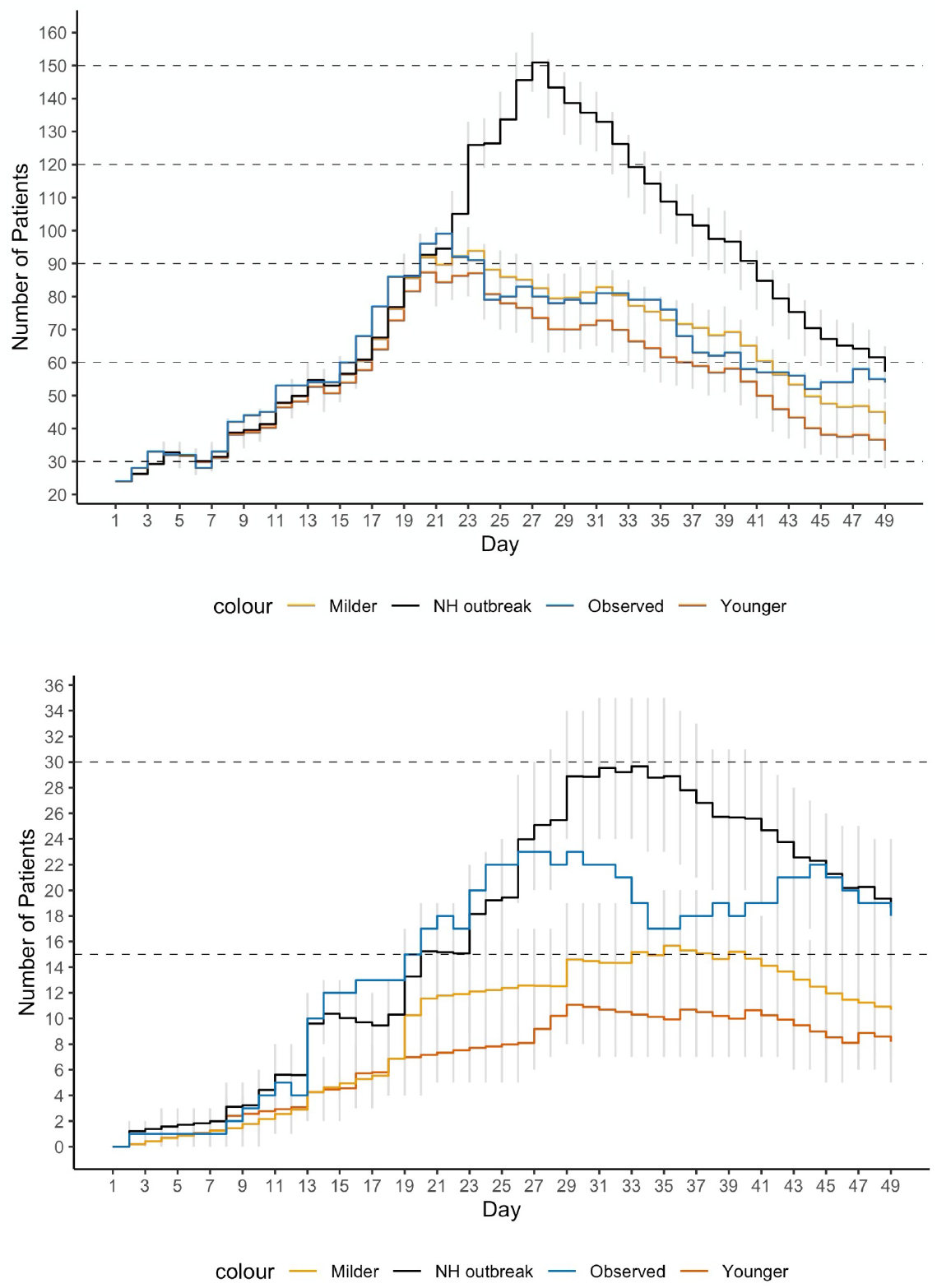
Prediction for hypothetical scenarios. Observed total hospitalized (top) and critical (bottom) patients, and predicted total number of hospitalized patients, under the following scenarios (i) “younger”: rate and state of incoming patients are the same as in Israel during the weeks from March 15th to May 2nd, but with patients in their 50s and 60s instead of 60+; (ii) “milder”: rate and age of incoming patients are the same as in Israel during the weeks from March 15th to May 2nd, but all patients incoming only in moderate and severe state, none at critical, and (iii) “Nursing home (NH) outbreak” where we assume that in addition to the arrival of patients as happened in Israel from March 15th to May 2nd there is a single week during which there are four times as many incoming patients aged 70+, arriving in various clinical states. Gray vertical lines are point-wise 10%-90% confidence predictions.

As an example of how these scenarios could be useful for resource planning, we consider the following use case for our model: COVID-19 patients are usually cared for in special wards. Our model can help planners assess when a new COVID-19 ward will need to open; towards that end, we add markers to Figure 3 indicating when total patient utilization passes multiples of 30, assuming that each COVID-19 ward can care for 30 patients. The intersection of the horizontal lines with the predicted utilization curve indicates at what date we estimate a new ward will need to be opened. Similarly, we add markers to show when critical patient utilization exceeds multiples of 15. In Figure S3 in the SM we show that the error for predicting the times when total hospital-bed utilization will hit such capacity thresholds is at most one day, and three days for critical-bed utilization.

## 4. Discussion

One of the distinctive characteristics of COVID-19 is the way health systems are overwhelmed by a large number of patients. For example, in Lombardy, Italy, ICU capacity reached its limit in early March, requiring urgent steps and outside assistance^2^ Similar events occurred in Madrid^12^, Wuhan^4^, the city of New York^3^, and other locations around the world.

We report here the development and validation of a multistate survival analysis model of patient clinical course throughout admission, discharge, and possibly death. Our model is based on the complete set of COVID-19 patients in Israel, tracked day-by-day; we note we had a very small number of patients with missing data (28 out of 2703).

We show that using very simple and easily available patient characteristics, a machine learning methodology based on a set of Cox regression models can accurately predict healthcare utilization for a given patient arrival process and can be used to simulate utilization under different patient influx scenarios. This can in turn be used to accurately plan resource allocation and the opening or closing of COVID-19 wards. We further provide an anonymized version of the dataset used to develop the model, a web-application for patient level predictions, and an R software package that can help planners fit a multistate model to their own data, or use the model we fit to the Israeli data.

Interestingly, we find that scenarios such as the arriving patients being much younger or in milder clinical state do not greatly affect total hospital utilization, possibly because some of these populations have longer hospitalization times; on the other hand, both scenarios affect critical-care bed utilization. We further observe that an eldercare nursing home outbreak scenario leads to substantially higher total utilization and critical care utilization, underscoring the need to protect these communities not only in terms of preventing mortality, but also from the point of view of lowering the strain on hospital resources.

Many models exist for predicting the dynamics of COVID-19 case numbers and numbers of hospitalized patients. These models are usually based on some variation of the basic susceptible-infected-recovered (SIR) model^13,14^, where the number of hospitalized patients are included as a component in the dynamic model. Both Hazard et al. ^15^ and Schmidt et al. ^16^ propose using a multistate model focusing only on patients admitted to the ICU in the most severe of clinical states, requiring extracorporeal membrane oxygenation (ECMO); the cohorts in these studies are small (77 and 83 patients, respectively), and are relevant only to the subset of COVID19 patients requiring ECMO treatment.

Our model differs from these models in several aspects: (i) Settings in which the chance of experiencing one event is altered by the occurrence of other events are known as competing and semi-competing risks, and caution is needed in analyzing such data. In semi-competing risks a subject can experience both a nonterminal and terminal event where the terminal event (e.g., death) censors the nonterminal event (e.g. being in a critical state), but not vice-versa. Patient clinical course data consist of competing and semi-competing risks. Ignoring (semi-) competing risks in time-to-event analyses can lead to substantially biased risk predictions^11,17^ In this work we use a multistate model as an excellent fit for the competing and semi-competing risks data of the COVID-19 patient hospitalization course. (ii) Heterogeneity within and between patients is important, as bed utilization is in part determined by a long tail of some patients who require significantly longer stays than others. (iii) Our model has the capability of modeling patients on an individual basis and takes into account how long each patient has already been in the hospital. Thus, our model will take into account the case-mix and heterogeneous histories of the patients currently hospitalized when making predictions about future utilization. We also emphasize that our model is different in its scope: we do not aim to model the spread of the disease and the number of future infections. Our focus is on estimating hospital-utilization under different patient arrival processes, while taking into account the load caused by currently hospitalized patients.

Another line of work related to ours are models for predicting outcomes for individual patients^18^ Viewed through this lens, our model is distinctive in two ways: it provides time-to-event (release, deterioration, death) predictions, and it is based on a very small number of covariates (age, sex, and one of three patient clinical states). For example, Liang et al. ^19^ report AUROC of 0·88 for predicting critical illness or death using ten covariates selected from 72 potential predictors. Bello-Chavolla et al. ^20^ report a concordance of 0·83 using seven covariates based mostly on comorbidities. We conjecture that the accuracy achieved by our model while using minimal, easily obtainable data as input might be explained by the fact that reported patient clinical states function, in a sense, as expert indicator variables summarizing more granular clinical measures and comorbidities.

Our model has several limitations. First, it is based on data from the first wave of patients in Israel. As treatment strategies and hospitalization policies differ over time and between health systems and hospitals, we cannot guarantee that LOS statistics will be the same across all locales and times. Thus, when possible we encourage planners to use the attached software package and fit it to their own hospitalization data. We will update the software package and app as more updated data will become available from the Israeli registry. A second limitation is that our model relies on estimation of the frequency and characteristics of future incoming patients. If arriving patient populations – both patient type and patient numbers – will differ significantly from the scenarios taken into account, the model’s predictions will be wrong. We thus recommend that planners evaluate multiple hypotheticals for incoming patients, testing for scenarios such as the ones we presented in the Results section above. A third limitation is that the model does not take into account patients’ comorbidities^21–23^ On the one hand, our model achieves good results while analyzing only a limited number of covariates as input; on the other hand, it is possible that using comorbidities could enhance the model’s performance. We also wish to point out that researchers with access to patient-level comorbidity data can easily incorporate it into a multistate model using the software we provide. A fourth limitation is that we used the patients’ clinical state as reported by the attending physician at the point of care. Although the Israeli MOH directed physicians to report clinical state using the definitions above, individual physicians and medical centers might not have adhered exactly to these guidelines. We note that despite this possible ambiguity, empirically we find that the clinical state as reported is indeed predictive for individual patients.

## 5. Conclusions

We found that a very small set of covariates (age, sex and patient being in one of three clinical states), along with a day-by-day tracking of patients’ clinical state, are enough for accurate predictions of mortality, length-of-hospitalization and critical illness. These accurate predictions enable us to build a tool that lets healthcare managers accurately plan resource allocation for COVID-19 patient care in the face of potentially large patient surges.

## Data Availability

An anonymized version of the data used for this study will be made available in the near future.

## Acknowledgments

We thank Dr. Amit Huppert and the biostatistics unit researchers at Gertner Institute for their insights and help in conducting this study. We thank Prof. Orly Manor for her valuable comments on the manuscript. We further thank Prof. Gadi Segal for insightful discussions. We wish to thank the Medical division and Information and Technologies division of the Israeli Ministry of Health for their efforts in the gathering and organization of the clinical data from all the Israeli hospitals. We thank the information and technologies staff of medical centers across Israel for building the data infrastructure needed for the collection of the data used in this study.

## Ethics committee approval

An exemption from institutional review board approval was determined by the Israeli Ministry of Health as part of an active epidemiological investigation, based on use of anonymous data only and no medical intervention.

## Authors contribution

MR contributed to acquisition of data, study conception and design, writing of the first draft, and critical revision of the manuscript and the supplementary material for important intellectual content. RG contributed to acquisition of data, statistical analysis, critical revision of the manuscript for important intellectual content. JS, ABA, IC contributed to programming required for the statistical data analysis. UG, SLT, AZ contributed to acquisition of data. YBL contributed to a critical revision of the manuscript for important intellectual content. DE contributed to study conception and design, critical revision of the manuscript for important intellectual content. MG contributed to study conception and design, statistical analysis and interpretation of the results, wrote the supplementary material, and critical revision of the manuscript for important intellectual content. US contributed to study conception and design, analysis and interpretation of data, writing of the first draft of the manuscript, and critical revision of the manuscript and of the supplementary material for important intellectual content.

## Supplementary Material

### S1 Models and Methods

#### S1.1 Introduction

The hospitalization course of each patient is described as a multi-state process, depicted in Figure S1. A patient enters the hospital at one of the following three clinical states: moderate, severe or critical. During the course of hospitalization a patient can move among the transient clinical states: Critical, denoted by *C*; Moderate or Severe, denoted by *M/S*; and Discharged, denoted by *Di*. Our multi-state model combines Moderate and Severe, during hospitalization, due to the small number of observed transitions from and to each of these states, separately. The state *Di* is considered as a transient state rather than a terminal state since frequently patients in a milder state were released from hospital to a dedicated quarantine for COVID-19 patients, with some later experiencing deterioration leading to re-hospitalization; in Figure S1 we see there were 102 transitions from state *Di* to state *M/S*. The terminal state Deceased is denoted by *De*.

Our multi-state model allows the following six transitions

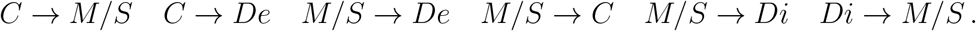

Three transitions were excluded from the model due to small sample size: The dataset includes 10 records of transition *C* → *Di*, no records of *Di* → *De*, and 2 records of *Di* → *C*. Hence, these three transitions were excluded from the multi-state model. Each possible transition is characterized by a transition-specific Cox proportional hazard model with an unspecified transition-specific baseline hazard function, λ0.,. and a transition-specific vector *β*.,. of regression coefficients. Specifically, for *t>* 0, the corresponding Cox proportional hazard functions are

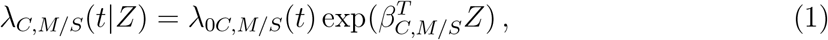

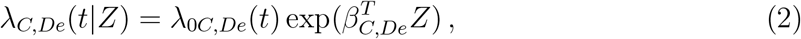

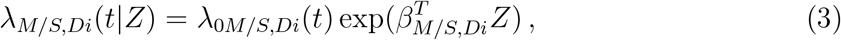

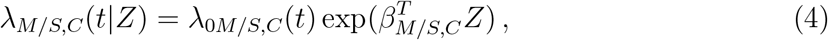

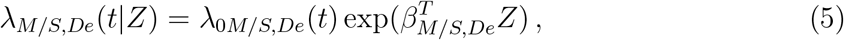

and

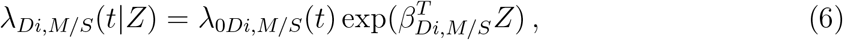

where *Z* is a vector of covariates, possibly with time-dependent covariates. For simplicity of notation, we use *Z* instead of *Z*(*t*) whenever confusion is unexpected. Although *Z* is shared by the six models above, it does not imply that identical covariates must be used in these models, since the regression coefficient vectors, *β*·,·, are transition dependent, and one can set any specific coefficient to 0 in order to exclude the corresponding covariate.

The covariates included in the Cox models were: age, sex, state at time of hospitalization (three categories: Moderate, Severe, or Critical), a binary variable equal to 1 if the patient previously was in a critical state and 0 otherwise, the cumulative number of days in hospital at entry time to the current state, and the interaction between age and each of the other covariates listed above. The models of transitions *M/S* → *De* and *Di* → *M/S* are slightly different due to only a few events with a critical clinical state at time of hospital admission and a few events with previous visits in critical state. Therefore, for these two models, the binary covariate taking the value of 1 if the patient previously visited the critical state is excluded, and the covariate for clinical state at time of hospitalization was redefined as a binary covariate indicating Moderate versus Severe/Critical.

#### S1.2 Estimation

Estimating the above hazard functions (1)–(6) involves several major issues (beside right censoring), which we describe below: multi-state process, left truncation, competing risks and recurrent events.

**Multi-state process:** We describe the hospitalization path of each patient as a multi-state process, starting at states *C* or *M/S*. Each patient may visits a transient state (*C*, *M/S* or *Di*) multiple times before reaching a terminal state (*De*).

**Left truncation:** Consider, for example, a patient who entered the hospital at state *M/S* and moved to state *C* at the 10th day of hospitalization. The contribution of such a patient to transitions from state *C* (back to *M/S* or to *De*) starts only at the 10th day of hospitalization. Hence, when estimating the model of a certain transition from origin state (e.g. *C*) to a target state (e.g. *De*), those who entered the origin state during the course of hospitalization are left-truncated by their entering day to that origin state.

**Figure S1:**
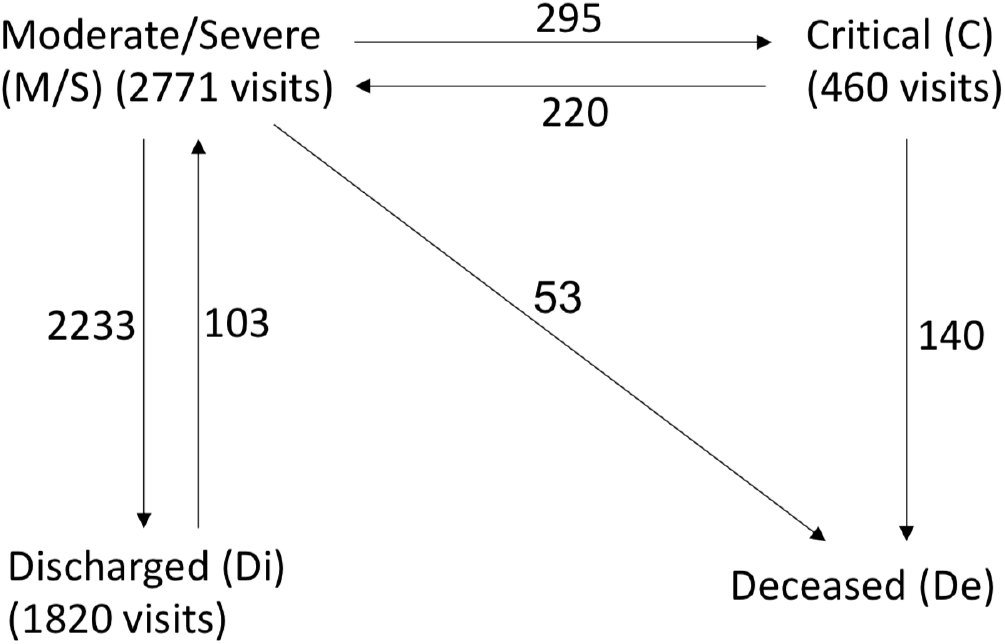
Multi-State Model: Data as of May 2, 2020, *n* = 2675. We model a COVID-19 patients disease course as moving between 4 possible clinical states: (i) moderate or severe, (ii) critical, (iii) discharged and (iv) deceased. We combined the two clinical states moderate and severe into a single model state due to statistical considerations; however, we keep a distinction between the two by a covariate indicating whether the patient first entered at mild/moderate clinical state or at a severe clinical state. Numbers next to arrows indicate number of observed transitions; each patient can make several clinical state transitions, and may visit a transient clinical state more than once. The dataset includes 10 records of transitions *C* → *Di*, no records of *Di* → *De*, and 2 records of *Di* → *C*. Hence, these three transitions were excluded from the multi-state model.

**Competing risks (competing events):** Given that a patient is, for example, at state *C*, there are two possible transitions: *C* → *M/S* and *C* → *De*. Since only one of these transitions can occur at each point in time, *M/S* and *De* are competing events in the sense that at each time point, the occurrence of one type of event will prevent the occurrence of the other. Similarly, given that a patient is at state *M/S*, the events *Di*, *C* and *De* are competing events.

**Recurrent events:** A patient may visit states *C*, *M/S* and *Di* multiple times. For example, 13 patients had the following hospitalization path

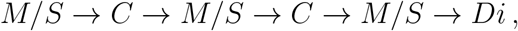

and 68 patients had

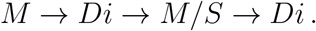

All the observed paths and their frequencies are provided in Table S1. When the event of interest can occur more than once in a patient, the events are termed recurrent events.

We overcame all the above challenges and provided consistent estimators (under mild regularity conditions) for the six Cox proportional hazard models. Specifically, by extending the approach of Andersen et al. (1991), it can be shown that maximizing the likelihood function in terms of the six Cox models can be done by maximizing the likelihood of each transition separately, while using the risk-set correction for dealing with left truncation (Klein & Moeschberger, 2006); treating competing transitions as right censoring (Kalbfleisch & Prentice, 2011); and adopting Andersen-Gill approach for dealing with recurrent events so the robust standard errors account for correlated outcomes within a patient (Andersen & Gill, 1982).

#### S1.3 Predictions

Based on our multi-state model, we accurately predict at the **patient level**, given age, sex and state at time of hospitalization, the following quantities:

1. The chance of in-hospital mortality (state *De*).
2. The chance of being at a critical state (state *C*).
3. The total length of stay (LOS) in hospital (not including time in a dedicated out-of-hospital quarantine).
4. The total length of stay in critical state (LOSCS).

The above quantities can be predicted at the first day of hospitalization and also during the course of hospitalization, while correctly taking into account the accumulated hospitalization history.

Weighted estimators of the area under the Receiver Operating Characteristic curve (AUROC) were used to evaluate death and critical-visit predictions for binary classifications (i.e. yes/no in-hospital mortality and yes/no previously visiting critical state). The weights eliminate the bias due to exclusion of censored observations, and were defined as 1 over the probability of being uncensored (Robins & Finkelstein, 2000). The weights were estimated by a Kaplan-Meier estimator of the censoring survival function. Weighted AUROC estimates were calculated by the R package WeightedROC (Hocking, 2020). Brier scores were used as measures of prediction accuracy.

Based on the above predictions, we go one step further and provide predictions at the **hospital level**, in the following manner:

1. **Snapshot:** Assume that at a given calendar day, we are given the current state and hospitalization history of all the COVID-19 patients currently at a specific hospital. Beyond predicting the above quantities for each patient, we also predict the total number of patients at the hospital, and at a critical clinical state in particular, for each day over the next 8 weeks. Namely, we provide predictions on the total occupancy on a calendar scale which are only due to the currently hospitalized patients.
2. **Arrival:** Given as input the arrival process of patients to the hospital at each day, including the number of arriving patients, their age, sex and state at time of hospitalization, we predict the total number of patients at hospital, and at critical state in particular, for each day of the next 8 weeks. Here we provide a prediction for the total occupancy on a calendar scale, for any possible hypothetical arrival scenario.
3. **Arrival plus Snapshot:** At a given calendar day, we are given the current state and hospitalization history of all the COVID-19 patients currently at the hospital along with an arrival process of the patients to be hospitalized starting the next day up to a pre-specified time period. Again, we predict the total number of patients at hospital, and at critical state in particular, on a calendar scale.

#### S1.4 Monte Carlo Estimator of Length of Stay

Since our hospitalization model consists of a multi-state model with recurring events (i.e. a patient can visit a transient state multiple times) the closed form marginal probabilities required for predictions are intractable. Instead we use a Monte Carlo (MC) approach for estimating all the required quantities listed above (Section S1.3).

Assume a prediction is desired for a new patient with baseline covariates (i.e. age, sex, and clinical state at time of hospitalization) denoted by *X*. The MC-based prediction procedure can be summarized as follows. Given *X*, sample a large number (e.g. 20,000) of hospitalization paths, and use these paths for estimating the required quantities. Specifically, the probability of death is estimated by the proportion of paths ended at state *De*; the probability of visiting state *C* is estimated by the proportion of paths visited state *C*; the expected total length of stay is estimated by the mean length of the paths (not including time at state *Di*); and the expected length of stay in state *C* is estimated by the mean time spent in state *C* over all the paths. The next subsection provides a detailed description of path sampling.

#### S1.5 Path Sampling - Technical Details

Let *J_C_* and *J_N_* denote the current and next states, respectively. Assume a patient entered the hospital at state *J_C_* = *j*^*^ with a vector of baseline covariates *X*. The goal is to provide a MC estimator of the length of hospitalization given *X* and *j*^*^ Let *Z*(*t*) be a time-dependent vector of covariates such that 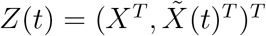, where 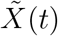 is a time-dependent vector of covariates that are known at the entrance to the new state. Details of 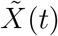 in our setting are provided at the end of Section S1.1. Assume *K_j*_* possible transitions from state *j*^*^ For each state *j*, *j* =1*,…, K_j*_*,

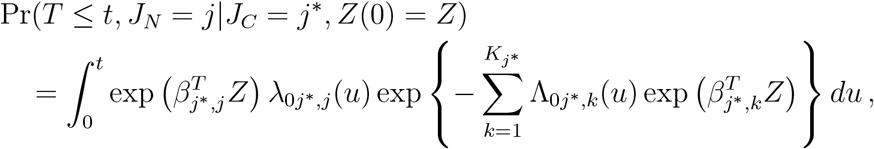

where *β*_j^*^,j_, λ_0j^*^,j_ and Λ_0j^*^,j_ are the vector of regression coefficients, the baseline hazard function and the cumulative baseline hazard function of transition *j*^*^ → *j*, respectively.

Then,

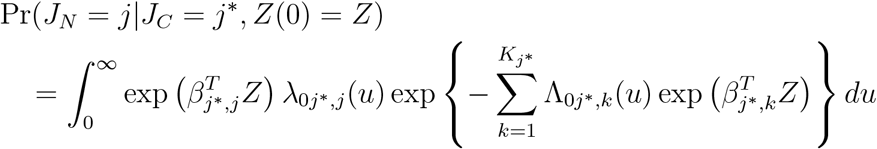

and

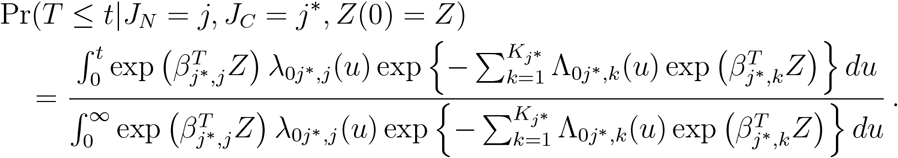

We start by describing the sampling procedure of the next state. Let *τ_j_*_*_*_,j_* be the largest observed event time of transition *j*^*^ → *j*. Then, the next state is sampled from a *K_j_*_*_ multinomial distribution with probabilities *p_j|j*,Z_* where, for *j* =1*,…,K_j_*_*_,

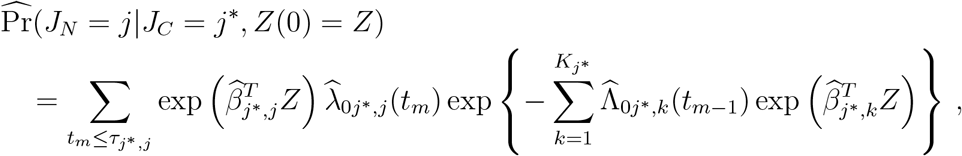

the summation is over the distinct observed event times of transition *j*^*^ → *j* and

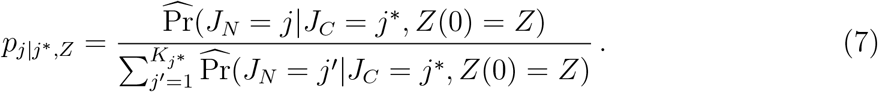

Once we sampled the next state, denoted by *j*^’^, the time to be spent at state *j*^*^ should be sampled based on

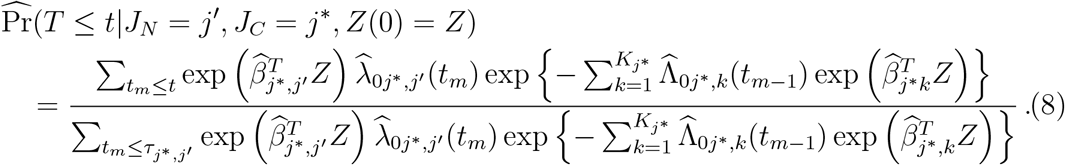

This could be done by sampling *U* ~ *Uniform*[0, 1], equating

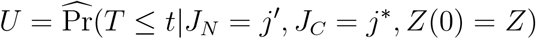

and solving for *t*. Denote the sampled time by *t*′ and update *Z*(*t*′). In case *j*′ = *De*, the sampling path ends here. Otherwise, the current state is updated to *J_C_* = *j*′, and the following state is sampled by *p_j_*|*_j′,Z_*, where for *j* =1*,…,K_j′_*

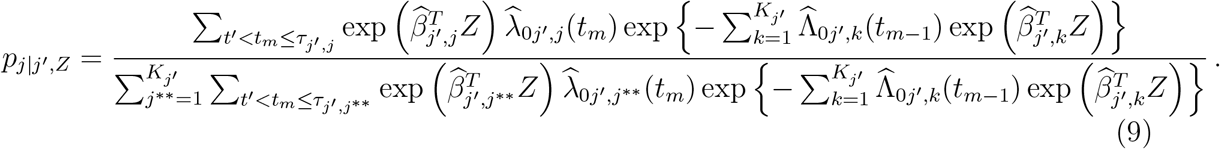

An exceptional state is *J_C_* = *Di*, where one can either move back to *M/S* or stay at *Di*. Given the new sampled state, denoted by 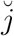, the time to be spent at *j*′ is sampled by

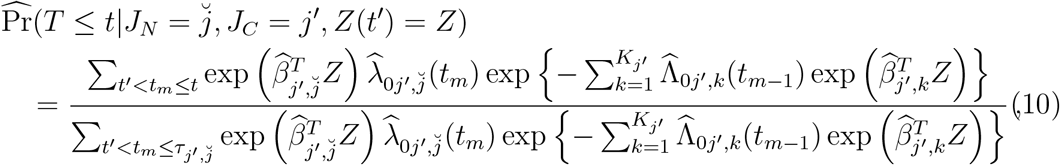

and then by solving for 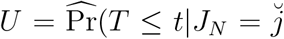*, J_C_* = *j*′*, Z*(*t*′)= *Z*). The sampled path is completed once state *De* is sampled or state *Di* is sampled and the next sampled state is again *Di*, or after sampling 9 states, whichever comes first. We set the maximum number of transitions to 9 as observed in our dataset (see eTableS1).

#### S1.6 Predictions at the Patient Level - Technical Details

We consider the following three types of predictions. Type A is prediction at time of hospitalization, Type B is prediction at the entrance to any new clinical state, and prediction Type C is done during the stay at a certain state. Let *Z*(*t*) consist of age, sex, clinical state at time of hospitalization, the cumulative number of days in hospital **up to the entrance to the current state**, an indicator variable equal to 1 if previously visited Critical state and 0 otherwise, and the interactions of each of these covariates with age.

**Type A**. Given age = *a*, sex = *s* (*s* is either 1 or 0) and state at time of hospitalization *j*^*^, the vector of covariates for prediction at time of hospitalization is given by

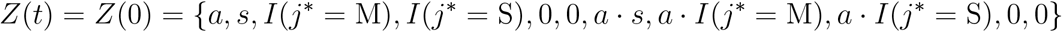

and Eq’s (7) and (8) provide the estimated probabilities of the next state *j* and the probability of transitioning by day *t* given the transition *j*^*^ → *j* and *Z*(0).

**Type B**. Given age = *a*, sex = *s*, state at hospitalization *j*^*^, the patient now entered state *j*′, the cumulative number of days in hospital up to the entrance to the current state equals *t*′, the vector of covariates for predictions is

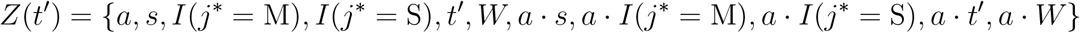

where *W* = 1 if previously visited *C*, and 0 otherwise. Hence, Eq’s (9) and (10) provide the estimated probabilities of the next state *j* and the probability of transitioning by day *t* given the transition *j*^*^ → *j*′ and *Z*(*t*′).

**Type C**. Given *Z*(*t*′) as in Type B, and given that the patient is at state *j*′ for already *d* days, the following are the updated predictions where we take into account the *d* days in current state but there is no change in the vector of covariates *Z*(*t*′). Specifically, the next state probability *p_j_*|*_j′,Z,d_*, *j* =1*,…,K_j′_*, is given by

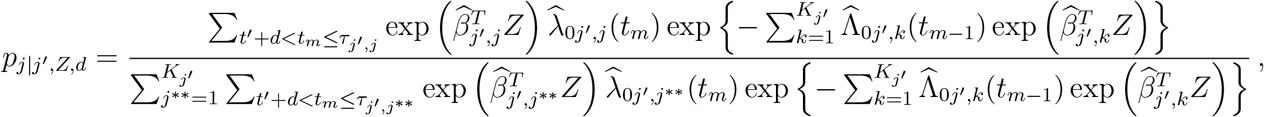

where summations are over the observed event times of the respective transition. Given the new sampled state, denoted by 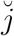, the time to be spent at *j*′ is predicted by

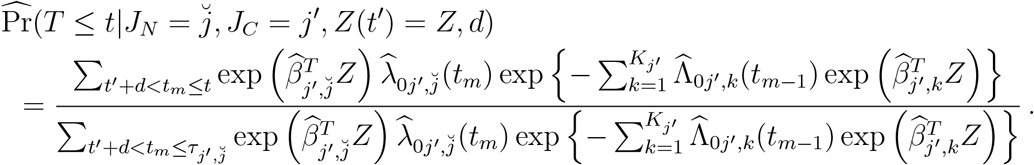

#### S1.7 Weighted Bootstrap Standard Error for Prediction at the Subject Level

Denote the total number of patients in the training data *n*. Our goal is estimating the standard error (SE) of predictions of a new patient with baseline covariate *X^o^*. The following is a weighted bootstrap procedure (Kosorok, 2007) for SE estimation of our proposed MC-based estimators:

1. Sample *n* weight values from an exponential distribution with mean 1, and assign a weight value to each patient in the training sample.
2. Estimate the six Cox PH models (1)–(6) with the weights sampled in Step 1.
3. Given *X^o^*, sample 20, 000 MC paths, and based on the 20,000 paths compute all the desired quantities according to Section S1.4.
4. Repeat Steps 1–3 *B* times.

The empirical variances of the *B* estimates are the weighted bootstrap SEs estimates. For example, the empirical variance of the *B* death probability estimates is the SE estimate of the estimated death probability of this new patient.

#### S1.8 Prediction at the Health-System Level

As described above, for prediction at the patient level, for any given vector of covariates one should run a large number of MC paths, summarize the MC runs and obtain the distributions of LOS in hospital, of LOSCS, etc. Predictions at the hospital level for Snapshot, Arrival or Snapshot plus Arrival (see Section S1.3 for details) require a different MC approach, as follows:

1. Sample one hospitalization path for each patient at the starting date for Snapshot patients and at the patient hospitalization date for Arrival patients.
2. Summarize the paths over all the patients during the predefined period. For example, count the number of patients in the hospital at each day.
3. Repeat Steps 1–2 above a large number of times (e.g. 10,000), and summarize over these repeats. For example, get the mean or various quantiles of the number of patients in the hospital at each day.

### S2 Results

Data analysis was conducted in R (R Core Team, 2020). Table S2 – Table S4 provide the regression coefficients of each of the transition models (1)–(6). The following are summaries of the various MC predictions.

#### S2.1 Prediction at the Patient Level

Table S5 provides death probabilities for various patient types defined by sex, age and state at time of hospitalization. Prediction is based on 20,000 MC paths for each patient type along with weighted-bootstrap 95% confidence intervals. As expected, death probability increases with age and with state severity at time of hospitalization. Table S6 gives the probability of being at critical state during hospitalization by patient types along with weighted-bootstrap 95% confidence intervals. The 10%, 25%, 50%, 75% and 90% quantiles of LOS in days and weighted-bootstrap standard errors, by patient type are given in Table S7. Quantiles of LOSCS in days, given being in critical state with weighted bootstrap standard errors are presented in Table S8 by patient type. Plots of the cumulative distribution function of LOS by patient types are provide in Figure S2. All the above results are based on 20,000 MC paths for the estimates of each patient type, and 500 weighted bootstrap samples, each with 20,000 MC paths, for the standard error estimates for each patient type.

The AUROC and Brier Score results, based on 8-fold cross validation, are presented in Table S10 (more details on the 8-fold cross validation study in the next section). The mean AUROC and Brier Scores estimates for death prediction over the eight held-out subsets are 0.955 (SE=0.035) and 0.043 (SE=0.011); the respective AUROC and Brier score of predicting critical clinical state visit are 0.880 (SE=0.040) and 0.049 (SE=0.013).

#### S2.2 Prediction at the Health System Level -Random Subset of Patients 8-fold Cross Validation

The entire dataset was randomly partitioned into 8 groups of about 330 patients each. Each time the model was trained with one group omitted (held-out), and our prediction tool was used to provide predictions for the patients of the held-out group. We demonstrate the results of one held-out random group in Figure S3 for the arrival process and two snapshots, at April 1st and 15th. Evidently, our prediction tool performs very well in terms of absolute error. Similar conclusions are obtained from Figure S4 which provides the summary over the 8 held-out groups. Tables S9 presents the mean absolute errors (MAE) of each held-out group and over the eight groups. MAE is defined as the mean of the absolute daily-based differences between observed and predicted number of patients; the means are over 64, 32 and 18 days, for the Arrival and the two Snapshot settings, respectively. “ALL” (in Tables S9) refers to MAE between the mean curves of the estimated and predicted curves of the 8 held-out random groups, and “Mean” (SE) are the means and SEs of the 8 MAEs. The results show excellent prediction performances with small mean MAE. In particular, 4.72 (SE=1.07) and 1.68 (SE=0.40) for LOS and LOSCS of Arrival; 3.15 (SE=1.20) and 1.47 (SE=0.56) for LOS and LOSCS of April 1st Snapshot; and 3.13 (SE=1.07) and 1.98 (SE=0.93) for LOS and LOSCS of April 15th Snapshot.

#### S2.3 Prediction at the Health System Level - Hospital Holdout

We further evaluate the model by training on a sample where patients from each hospital in turn are held-out and not included in the training dataset. Figure S5 and Figure S6 demonstrate our ability to predict for the two hospitals with the largest patient population in our sample, hospitals H5 and H7 (we were asked to avoid identifying the hospital names). Evidently, load predictions for H5 are satisfactory, but not so for H7. It shows that when using our model for predicting at the hospital level, it is preferable to include patients from the predicted hospital in the training data thus avoiding possible hospital-specific biases. Table S11 and Table S12 show the mean absolute error, ROC AUC and Brier Scores for predicting death and visiting critical state for each hospital. In some hospitals, the number of deceased or number of patients with visits at Critical are very small, so results should be interpreted with caution.

#### S2.4 Prediction at the Health System Level -Hypothetical Scenarios

Assume we would like to predict the hospitalized load in a certain health-care system in a given period of time using the Snapshot plus Arrival approach (see Section S1.3 for details). Three hypothetical scenarios were constructed based on the observed arrival process of a random hold-out sample of size 330 (specifically, we used the first random subset described in Section S2.2 and Figure S3):

1. **“Younger”**: patients of age 60 and above within the held-out sample were replaced by patients of ages between 40–50 with probability 2/3, and between 50–60 with probability 1/3. Given the age group of a new patient, the specific age, sex and clinical state at time of hospitalization were sampled based on the distribution of Israeli COVID-19 patients.
2. **“Milder”**: patients at the critical clinical state at time of hospitalization were left at the critical state or replaced by Severe or Moderate patients, each with probabilities 1/3. Given the state at time of hospitalization of a new patient, the specific age and sex were sampled based on the distribution of Israeli COVID-19 patients.
3. **“Elder Care Nursing Home Outbreak”**: On the 5th week from the beginning of the arrival process, and only for this week, the number of patients at age 70 and above was multiplied by four. Age and sex of the new patients were sampled based on the Israeli population distribution, and the state at time of hospitalization was sampled based on the distribution of Israeli COVID-19 patients.

The model was trained by the data not including the random held-out sample, and prediction was performed with 10,000 MC paths as described in Section S1.8. The results are presented in Figure S7 as well as in Figure 2 of the main paper. Table S13 (same as Table 3 of main paper) shows the number of observed death based on the actual data of the held-out sample, versus the expected number of deaths for each of the above hypothetical scenarios. The high similarity between the observed and expected number of deaths demonstrate that our proposed model is well calibrated. Under the “Younger” scenario the number of deaths decreases dramatically, the decrease is moderate under the “Milder” scenario, and as expected, an outbreak at elder care nursing home yields a substantial increase in number of deaths.

**Table S1:** Summary of the observed hospitalization course (observed paths): Di -Discharged, M/S -Moderate/Severe, C -Critical, De -Deceased. A patient enters the hospital at a moderate, severe or critical clinical state, and can move among the transient clinical states during the course of hospitalization. The longest observed path consists of 9 transitions.

|  | path | frequency |
| --- | --- | --- |
| 1 | M/S | 148 |
| 2 | M/S Di | 1977 |
| 3 | M/S Di M/S | 19 |
| 4 | M/S Di M/S Di | 68 |
| 5 | M/S Di M/S Di M/S | 1 |
| 6 | M/S Di M/S Di M/S Di | 5 |
| 7 | M/S Di M/S Di M/S Di M/S Di M/S Di | 1 |
| 8 | M/S Di M/S C | 2 |
| 9 | M/S Di M/S De | 2 |
| 10 | M/S Di C | 1 |
| 11 | M/S Di C M/S Di | 1 |
| 12 | M/S C | 49 |
| 13 | M/S C Di | 4 |
| 14 | M/S C M/S | 25 |
| 15 | M/S C M/S Di | 61 |
| 16 | M/S C M/S Di M/S Di | 1 |
| 17 | M/S C M/S C | 8 |
| 18 | M/S C M/S C M/S | 4 |
| 19 | M/S C M/S C M/S Di | 13 |
| 20 | M/S C M/S C M/S Di M/S | 1 |
| 21 | M/S C M/S C M/S C | 1 |
| 22 | M/S C M/S C M/S C M/S | 1 |
| 23 | M/S C M/S C M/S C M/S C | 1 |
| 24 | M/S C M/S C M/S C De | 1 |
| 25 | M/S C M/S C De | 3 |
| 26 | M/S C M/S De | 2 |
| 27 | M/S C De | 64 |
| 28 | M/S De | 44 |
| 29 | C | 42 |
| 30 | C Di | 6 |
| 31 | C M/S | 12 |
| 32 | C M/S Di | 33 |
| 33 | C M/S Di M/S | 1 |
| 34 | C M/S C | 3 |
| 35 | C M/S C M/S | 2 |
| 36 | C M/S C M/S Di | 6 |
| 37 | C M/S C M/S C | 1 |
| 38 | C M/S C M/S C M/S | 2 |
| 39 | C M/S C M/S C M/S Di | 2 |
| 40 | C M/S C M/S C M/S C | 1 |
| 41 | C M/S C M/S C De | 2 |
| 42 | C M/S C M/S De | 1 |
| 43 | C M/S C De | 3 |
| 44 | C M/S De | 4 |
| 45 | C De | 74 |

**Table S2:** Results of Cox survival analysis models of transitions from Critical state: STH: state at time of hospitalization (Moderate/Severe/Critical); HistCrt equals 1 if visited Critical state previously; Cum. Days is the number of days in hospital at entry to current state. The reported values are the regression coefficients (robust SE, p-value if smaller than 0.05). Results are based on 460 state visits (a patient can visit the state multiple times). Next states: 140 to Deceased state, 220 to Moderate/Severe state, and 10 to Discharged state.

|  | Transition to Deceased<br>estimate (SE, pvalue) | Transition to Moderate/Severe<br>estimate (SE, pvalue) |
| --- | --- | --- |
| Age | 0.022 (0.023) | 0.006 (0.009) |
| Sex (Male) | -3.156 (1.329, 0.018) | 0.278 (0.614) |
| STH (Severe) | -0.799 (1.841) | -0.471 (0.775) |
| STH (Critical) | -1.410 (1.761) | 0.543 (0.758) |
| Cum. Days | 0.059 (0.253) | 0.057 (0.078) |
| HistCrt (yes) | 3.499 (3.921) | 3.180 (1.451, 0.028) |
| Age:sex (Male) | 0.037 (0.017, 0.028) | -0.011 (0.009) |
| Age:STH (Severe) | 0.012 (0.024) | 0.006 (0.011) |
| Age:STH (Critical) | 0.021 (0.022) | -0.011 (0.011) |
| Age:Cum. Days | -0.0008 (0.003) | -0.0008 (0.001) |
| Age:HistCrt (yes) | -0.055 (0.046) | -0.030 (0.019) |

**Table S3:** Results of Cox survival analysis models of transitions from Moderate/Severe state: STH: state at time of hospitalization (Moderate/Severe/Critical); HistCrt equals 1 if visited Critical state previously; Cum. Days is the number of days in hospital at entry to current state. The reported values are the regression coefficients (robust SE, p-value if smaller than 0.05). Results are based on 2771 state visits (a patient can visit in the state multiple times). Next states: 53 to Deceased state, 295 to Critical state, and 2233 to Discharged state. Some covariates are not used in some of the models, see S1.1.

|  | Transition to Deceased<br>estimate (SE, pvalue) | Transition to Critical<br>estimate (SE, pvalue) | Transition to Discharged<br>estimate (SE, pvalue) |
| --- | --- | --- | --- |
| Age | 0.103 (0.026, 0.0000) | 0.029 (0.006, 0.000) | -0.015 (0.002, 0.000) |
| Sex (Male) | -3.039 (2.351) | 0.045 (0.531) | -0.307 (0.145, 0.034) |
| STH (Severe/Critical) | 4.278 (2.544) | - | - |
| STH (Severe) | - | 1.945 (0.589, 0.001) | 0.205 (0.186) |
| STH (Critical) | - | 1.275 (1.710) | 0.598 (0.899) |
| Cum. Days | -0.872 (0.532) | 0.010 (0.070) | -0.010 (0.027) |
| HistCrt (yes) | - | 1.643 (1.301) | -0.183 (0.660) |
| Age:sex (Male) | 0.037 (0.028) | 0.005 (0.007) | 0.005 (0.002, 0.028) |
| Age:STH(Severe/Critical) | -0.037 (0.030) | - | - |
| Age:STH (Severe) | - | -0.010 (0.008) | -0.009 (0.003, 0.002) |
| Age:STH (Critical) | - | -0.003 (0.0M/S) | -0.0132 (0.014) |
| Age:Cum. Days | 0.010 (0.006) | 0.000 (0.0009) | 0.0002 (0.0004) |
| Age:HistCrt (yes) | - | -0.016 (0.018) | -0.003 (0.010) |

**Table S4:** Results of Cox survival analysis models of transition from Discharged state to Moderate/Severe state: STH: state at time of hospitalization (Moderate vs. Severe/Critical); Cum. Days is the number of days in hospital at entry to current state. The reported values are the regression coefficients (robust SE, p-value if smaller than 0.05). Results are based on 1820 state visits (a patient can visit in the state multiple times). Next states: 0 at Deceased state, 2 at Critical state, and 103 at Moderate/Severe state.

|  | estimate (SE, pvalue) |
| --- | --- |
| Age | 0.043 (0.010, 0.0000) |
| Sex (Male) | 0.294 (0.763) |
| STH (Severe/Critical) | 0.648 (1.525) |
| Cum. Days | 0.287 (0.081, 0.0004) |
| Age:sex (Male) | 0.0017 (0.012) |
| Age:STH (Severe/Critical) | 0.007 (0.022) |
| Age:Cum. Days | 0.003 (0.001, 0.0099) |

**Table S5:** Death probability by patient type (state at time of hospitalization, age, sex) based on 20,000 MC paths for each patient type (weighted bootstrap 95% confidence interval).

| Patient Type | Male | Female |
| --- | --- | --- |
| Moderate, 15 | 0.000 (0.000,0.002) | 0.001 (0.000,0.004) |
| Moderate, 25 | 0.001 (0.000,0.003) | 0.002 (0.000,0.006) |
| Moderate, 35 | 0.002 (0.000,0.005) | 0.004 (0.000,0.009) |
| Moderate, 45 | 0.003 (0.000,0.007) | 0.007 (0.000,0.014) |
| Moderate, 55 | 0.009 (0.00,0.02) | 0.013 (0.00,0.02) |
| Moderate, 65 | 0.022 (0.01,0.03) | 0.026 (0.01,0.04) |
| Moderate, 75 | 0.058 (0.03,0.09) | 0.053 (0.03,0.08) |
| Moderate, 85 | 0.151 (0.09,0.21) | 0.122 (0.08,0.17) |
| Severe, 15 | 0.005 (0.00,0.01) | 0.009 (0.00,0.03) |
| Severe, 25 | 0.010 (0.00,0.03) | 0.017 (0.00,0.05) |
| Severe, 35 | 0.016 (0.00,0.04) | 0.027 (0.00,0.06) |
| Severe, 45 | 0.023 (0.00,0.06) | 0.046 (0.00,0.09) |
| Severe, 55 | 0.043 (0.00,0.09) | 0.075 (0.01,0.14) |
| Severe, 65 | 0.096 (0.04,0.15) | 0.126 (0.04,0.21) |
| Severe, 75 | 0.214 (0.15,0.28) | 0.217 (0.13,0.31) |
| Severe, 85 | 0.443 (0.32,0.56) | 0.389 (0.29,0.49) |
| Critical, 15 | 0.023 (0.00,0.11) | 0.061 (0.00,0.22) |
| Critical, 25 | 0.043 (0.00,0.15) | 0.099 (0.00,0.28) |
| Critical, 35 | 0.069 (0.00,0.19) | 0.153 (0.00,0.34) |
| Critical, 45 | 0.083 (0.00,0.22) | 0.224 (0.00,0.42) |
| Critical, 55 | 0.153 (0.04,0.27) | 0.299 (0.11,0.49) |
| Critical, 65 | 0.299 (0.18,0.42) | 0.414 (0.26,0.57) |
| Critical, 75 | 0.546 (0.43,0.66) | 0.549 (0.43,0.67) |
| Critical, 85 | 0.829 (0.74,0.91) | 0.754 (0.65,0.86) |

**Table S6:**
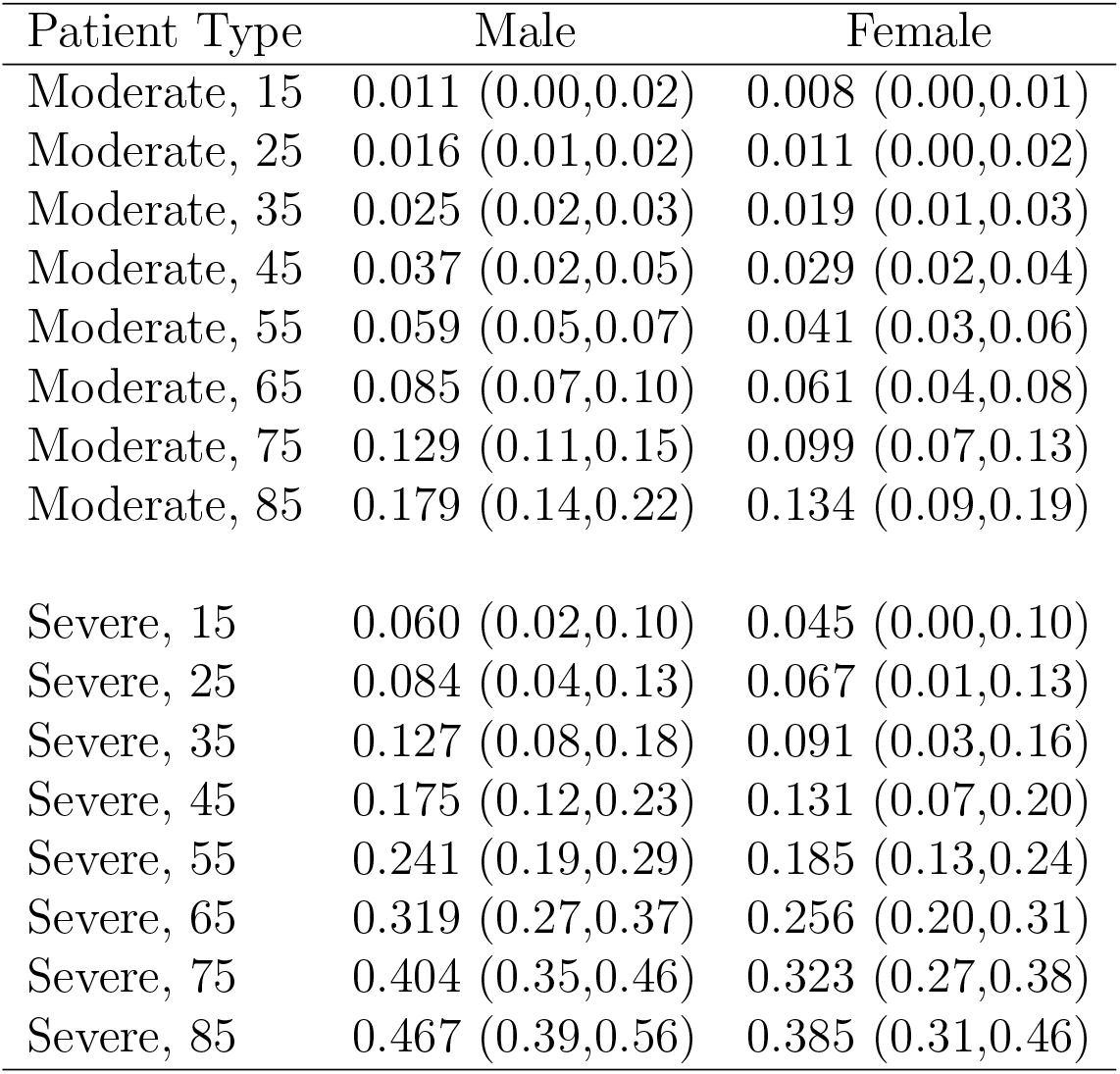
The probability of being at critical state during hospitalization by patient type (state at time of hospitalization, age, sex) based on 20,000 MC paths for each type (weighted bootstrap 95% confidence interval).

| Patient Type | Male | Female |
| --- | --- | --- |
| Moderate, 15 | 0.011 (0.00,0.02) | 0.008 (0.00,0.01) |
| Moderate, 25 | 0.016 (0.01,0.02) | 0.011 (0.00,0.02) |
| Moderate, 35 | 0.025 (0.02,0.03) | 0.019 (0.01,0.03) |
| Moderate, 45 | 0.037 (0.02,0.05) | 0.029 (0.02,0.04) |
| Moderate, 55 | 0.059 (0.05,0.07) | 0.041 (0.03,0.06) |
| Moderate, 65 | 0.085 (0.07,0.10) | 0.061 (0.04,0.08) |
| Moderate, 75 | 0.129 (0.11,0.15) | 0.099 (0.07,0.13) |
| Moderate, 85 | 0.179 (0.14,0.22) | 0.134 (0.09,0.19) |
| Severe, 15 | 0.060 (0.02,0.10) | 0.045 (0.00,0.10) |
| Severe, 25 | 0.084 (0.04,0.13) | 0.067 (0.01,0.13) |
| Severe, 35 | 0.127 (0.08,0.18) | 0.091 (0.03,0.16) |
| Severe, 45 | 0.175 (0.12,0.23) | 0.131 (0.07,0.20) |
| Severe, 55 | 0.241 (0.19,0.29) | 0.185 (0.13,0.24) |
| Severe, 65 | 0.319 (0.27,0.37) | 0.256 (0.20,0.31) |
| Severe, 75 | 0.404 (0.35,0.46) | 0.323 (0.27,0.38) |
| Severe, 85 | 0.467 (0.39,0.56) | 0.385 (0.31,0.46) |

**Table S7:** Quantiles of length of stay in days, by patient type (sex, age and state at time of hospitalization), based on 20,000 MC paths for each patient type (weighted bootstrap standard error).

| Patient Type | 10% | 25% | 50% | 75% | 90% |
| --- | --- | --- | --- | --- | --- |
| Male, 15, Moderate | 1 (0.37) | 3 (0.37) | 5 (0.37) | 8 (0.53) | 13 (0.92) |
| Male, 25, Moderate | 2 (0.49) | 3 (0.00) | 5 (0.45) | 9 (0.47) | 14 (0.88) |
| Male, 35, Moderate | 2 (0.41) | 3 (0.00) | 6 (0.19) | 10 (0.48) | 16 (0.95) |
| Male, 45, Moderate | 2 (0.01) | 3 (0.17) | 6 (0.19) | 11 (0.49) | 18 (0.97) |
| Male, 55, Moderate | 2 (0.00) | 4 (0.33) | 7 (0.17) | 12 (0.58) | 21 (0.69) |
| Male, 65, Moderate | 2 (0.00) | 4 (0.00) | 7 (0.50) | 13 (0.52) | 24 (1.06) |
| Male, 75, Moderate | 2 (0.00) | 4 (0.48) | 8 (0.50) | 15 (0.66) | 28 (1.44) |
| Male, 85, Moderate | 2 (0.32) | 5 (0.49) | 8 (0.54) | 16 (0.94) | 29 (2.05) |
| Male, 15, Severe | 1 (0.28) | 3 (0.50) | 5 (0.58) | 8 (0.92) | 14 (2.49) |
| Male, 25, Severe | 1 (0.50) | 3 (0.20) | 6 (0.56) | 10 (1.07) | 18 (3.14) |
| Male, 35, Severe | 2 (0.29) | 3 (0.39) | 6 (0.52) | 12 (1.29) | 25 (3.54) |
| Male, 45, Severe | 2 (0.00) | 4 (0.27) | 8 (0.56) | 16 (1.59) | 36 (4.26) |
| Male, 55, Severe | 2 (0.00) | 4 (0.49) | 9 (0.69) | 21 (1.45) | 42 (2.76) |
| Male, 65, Severe | 2 (0.49) | 5 (0.46) | 11 (0.88) | 26 (2.01) | 45 (1.62) |
| Male, 75, Severe | 3 (0.10) | 6 (0.36) | 12 (0.10) | 28 (2.08) | 47 (1.75) |
| Male, 85, Severe | 3 (0.26) | 6 (0.47) | 10 (0.10) | 23 (2.77) | 42 (5.89) |
| Male, 15, Critical | 6 (1.21) | 9 (2.22) | 16 (4.09) | 26 (6.87) | 41 (6.60) |
| Male, 25, Critical | 6 (1.16) | 11 (2.13) | 19 (3.78) | 31 (6.34) | 42 (4.79) |
| Male, 35, Critical | 7 (1.05) | 13 (1.92) | 22 (3.39) | 37 (5.17) | 45 (3.03) |
| Male, 45, Critical | 8 (0.92) | 14 (1.79) | 25 (3.04) | 42 (3.51) | 47 (2.08) |
| Male, 55, Critical | 8 (0.95) | 15 (1.78) | 28 (2.49) | 44 (2.55) | 48 (1.65) |
| Male, 65, Critical | 6 (0.69) | 13 (1.58) | 26 (2.39) | 45 (1.79) | 51 (1.76) |
| Male, 75, Critical | 3 (0.79) | 8 (0.86) | 20 (2.08) | 40 (4.71) | 48 (1.85) |
| Male, 85, Critical | 2 (0.47) | 4 (0.83) | 9 (1.25) | 21 (3.09) | 37 (5.84) |
| Female, 15, Moderate | 1 (0.00) | 2 (0.20) | 4 (0.24) | 7 (0.52) | 10 (0.87) |
| Female, 25, Moderate | 1 (0.10) | 2 (0.50) | 5 (0.50) | 8 (0.54) | 12 (0.88) |
| Female, 35, Moderate | 1 (0.40) | 3 (0.00) | 5 (0.34) | 9 (0.55) | 14 (0.86) |
| Female, 45, Moderate | 2 (0.31) | 3 (0.00) | 6 (0.17) | 10 (0.50) | 17 (0.97) |
| Female, 55, Moderate | 2 (0.00) | 3 (0.50) | 7 (0.37) | 12 (0.49) | 20 (1.04) |
| Female, 65, Moderate | 2 (0.00) | 4 (0.00) | 8 (0.46) | 13 (0.49) | 22 (1.17) |
| Female, 75, Moderate | 2 (0.22) | 4 (0.49) | 9 (0.49) | 15 (0.78) | 26 (1.52) |
| Female, 85, Moderate | 3 (0.39) | 5 (0.37) | 9 (0.51) | 17 (1.15) | 29 (1.85) |
| Female, 15, Severe | 1 (0.00) | 2 (0.22) | 4 (0.48) | 7 (0.84) | 11 (1.92) |
| Female, 25, Severe | 1 (0.20) | 3 (0.49) | 5 (0.53) | 8 (0.87) | 14 (2.34) |
| Female, 35, Severe | 2 (0.47) | 3 (0.00) | 6 (0.49) | 10 (1.07) | 20 (2.54) |
| Female, 45, Severe | 2 (0.10) | 4 (0.46) | 7 (0.47) | 13 (1.10) | 25 (2.59) |
| Female, 55, Severe | 2 (0.00) | 4 (0.17) | 8 (0.54) | 17 (1.22) | 31 (2.54) |
| Female, 65, Severe | 3 (0.49) | 5 (0.33) | 10 (0.62) | 21 (1.34) | 39 (3.14) |
| Female, 75, Severe | 3 (0.14) | 6 (0.36) | 11 (0.96) | 24 (1.89) | 42 (3.29) |
| Female, 85, Severe | 3 (0.50) | 6 (0.46) | 12 (1.20) | 25 (2.75) | 37 (4.90) |
| Female, 15, Critical | 5 (1.02) | 9 (1.24) | 15 (2.38) | 26 (4.63) | 39 (7.74) |
| Female, 25, Critical | 5 (1.13) | 9 (2.17) | 16 (4.13) | 27 (6.97) | 42 (7.02) |
| Female, 35, Critical | 6 (1.00) | 10 (1.94) | 18 (2.41) | 30 (3.59) | 42 (6.05) |
| Female, 45, Critical | 5 (1.10) | 10 (1.83) | 19 (3.17) | 32 (5.41) | 44 (3.60) |
| Female, 55, Critical | 5 (1.02) | 9 (1.65) | 20 (2.41) | 34 (3.97) | 45 (1.91) |
| Female, 65, Critical | 4 (1.03) | 8 (1.55) | 18 (2.73) | 34 (3.96) | 45 (1.87) |
| Female, 75, Critical | 3 (0.64) | 6 (1.07) | 15 (2.91) | 31 (4.23) | 47 (1.86) |
| Female, 85, Critical | 2 (0.68) | 5 (0.91) | 9 (1.86) | 24 (4.77) | 42 (5.62) |

**Table S8:** Quantiles of length of stay in critical state in days, given being in critical, by patient type (sex, age and state at time of hospitalization), based on 20,000 MC paths for each patient type (weighted bootstrap standard error).

| Patient Type | 10% | 25% | 50% | 75% | 90% |
| --- | --- | --- | --- | --- | --- |
| Male, 15, Moderate | 2 (0.69) | 5 (1.43) | 10 (2.66) | 19 (4.85) | 31 (6.48) |
| Male, 25, Moderate | 2 (0.64) | 5 (1.28) | 10 (2.36) | 20 (4.25) | 32 (4.91) |
| Male, 35, Moderate | 2 (0.56) | 5 (1.04) | 10 (1.96) | 21 (3.34) | 30 (4.10) |
| Male, 45, Moderate | 2 (0.49) | 6 (0.92) | 12 (1.83) | 22 (2.78) | 32 (3.11) |
| Male, 55, Moderate | 3 (0.49) | 5 (0.81) | 11 (1.24) | 22 (1.84) | 33 (2.31) |
| Male, 65, Moderate | 2 (0.44) | 5 (0.70) | 12 (1.21) | 21 (1.83) | 32 (1.94) |
| Male, 75, Moderate | 2 (0.14) | 4 (0.56) | 11 (1.15) | 20 (1.69) | 30 (2.50) |
| Male, 85, Moderate | 2 (0.48) | 4 (0.67) | 9 (1.45) | 17 (2.37) | 27 (3.07) |
| Male, 15, Severe | 3 (1.07) | 7 (2.37) | 14 (4.81) | 26 (6.84) | 36 (6.71) |
| Male, 25, Severe | 3 (0.95) | 6 (2.03) | 14 (3.92) | 25 (5.67) | 37 (5.47) |
| Male, 35, Severe | 3 (0.79) | 7 (1.78) | 14 (3.15) | 26 (4.55) | 37 (4.11) |
| Male, 45, Severe | 3 (0.63) | 7 (1.41) | 15 (2.46) | 26 (3.40) | 36 (3.07) |
| Male, 55, Severe | 3 (0.56) | 7 (1.87) | 15 (1.72) | 25 (2.45) | 34 (2.25) |
| Male, 65, Severe | 3 (0.40) | 6 (0.98) | 14 (1.49) | 24 (2.17) | 33 (2.13) |
| Male, 75, Severe | 3 (0.50) | 6 (0.76) | 13 (1.27) | 23 (1.75) | 31 (2.01) |
| Male, 85, Severe | 2 (0.50) | 4 (0.69) | 10 (1.68) | 19 (2.48) | 28 (2.68) |
| Male, 15, Critical | 2 (0.62) | 5 (1.45) | 11 (2.96) | 19 (4.71) | 30 (6.37) |
| Male, 25, Critical | 3 (0.64) | 6 (1.38) | 12 (2.55) | 23 (4.09) | 33 (5.56) |
| Male, 35, Critical | 3 (0.61) | 7 (1.24) | 14 (2.26) | 25 (3.28) | 37 (4.35) |
| Male, 45, Critical | 4 (0.55) | 8 (0.96) | 16 (1.66) | 27 (2.73) | 39 (3.23) |
| Male, 55, Critical | 4 (0.63) | 8 (1.04) | 17 (1.45) | 29 (2.00) | 40 (2.67) |
| Male, 65, Critical | 4 (0.62) | 8 (0.88) | 17 (1.29) | 30 (2.13) | 40 (3.51) |
| Male, 75, Critical | 3 (0.48) | 6 (0.63) | 14 (1.44) | 26 (1.99) | 37 (2.85) |
| Male, 85, Critical | 2 (0.43) | 4 (0.73) | 8 (0.90) | 18 (2.33) | 30 (2.71) |
| Female, 15, Moderate | 1 (0.85) | 4 (1.50) | 10 (3.69) | 21 (5.66) | 28 (7.07) |
| Female, 25, Moderate | 2 (0.66) | 4 (1.36) | 9 (3.08) | 15 (5.04) | 22 (6.35) |
| Female, 35, Moderate | 2 (0.52) | 4 (0.98) | 9 (2.16) | 16 (3.81) | 25 (5.21) |
| Female, 45, Moderate | 1 (0.57) | 3 (0.70) | 8 (1.63) | 16 (2.84) | 24 (4.02) |
| Female, 55, Moderate | 2 (0.46) | 3 (0.43) | 7 (1.09) | 14 (1.74) | 24 (2.52) |
| Female, 65, Moderate | 1 (0.47) | 3 (0.26) | 7 (0.84) | 13 (1.37) | 21 (1.77) |
| Female, 75, Moderate | 1 (0.48) | 3 (0.22) | 7 (0.78) | 13 (1.27) | 22 (1.72) |
| Female, 85, Moderate | 1 (0.43) | 3 (0.24) | 7 (0.92) | 13 (1.48) | 21 (2.12) |
| Female, 15, Severe | 3 (1.06) | 7 (2.31) | 14 (4.52) | 26 (6.50) | 35 (6.46) |
| Female, 25, Severe | 2 (0.86) | 5 (1.91) | 12 (3.83) | 24 (5.64) | 35 (5.89) |
| Female, 35, Severe | 2 (0.74) | 5 (1.56) | 11 (2.98) | 22 (4.73) | 32 (5.11) |
| Female, 45, Severe | 2 (0.55) | 5 (1.13) | 11 (2.34) | 20 (3.48) | 30 (4.28) |
| Female, 55, Severe | 2 (0.33) | 4 (0.67) | 9 (1.32) | 17 (2.10) | 27 (2.77) |
| Female, 65, Severe | 2 (0.32) | 4 (0.66) | 9 (1.01) | 16 (1.58) | 25 (2.17) |
| Female, 75, Severe | 2 (0.30) | 4 (0.53) | 8 (0.98) | 15 (1.50) | 23 (2.01) |
| Female, 85, Severe | 1 (0.47) | 3 (0.35) | 7 (1.14) | 14 (1.64) | 22 (2.21) |
| Female, 15, Critical | 2 (0.80) | 5 (1.80) | 11 (3.68) | 20 (6.36) | 32 (7.86) |
| Female, 25, Critical | 2 (0.68) | 5 (1.64) | 11 (3.15) | 21 (5.45) | 32 (6.91) |
| Female, 35, Critical | 3 (0.66) | 5 (1.34) | 11 (2.48) | 21 (4.39) | 33 (5.62) |
| Female, 45, Critical | 3 (0.60) | 5 (0.99) | 11 (1.99) | 21 (3.48) | 32 (4.56) |
| Female, 55, Critical | 3 (0.55) | 5 (0.75) | 11 (1.46) | 20 (2.47) | 31 (3.02) |
| Female, 65, Critical | 2 (0.54) | 5 (0.70) | 10 (1.33) | 20 (2.18) | 31 (2.89) |
| Female, 75, Critical | 2 (0.54) | 5 (0.55) | 9 (1.07) | 18 (2.07) | 28 (2.88) |
| Female, 85, Critical | 2 (0.43) | 4 (0.60) | 7 (0.84) | 15 (2.34) | 25 (3.04) |

**Table S9:** Random subset of patients holdout: Mean absolute error (MAE) for predicting number of hospitalized and number of critical patients over each held-out group and over the eight random groups together under the evaluation setups described in subsection S1.3.

| Group Number | Arrival All | Arrival Critical | Snapshot Apr 1st, All | Snapshot Apr 1st, Critical | Snapshot Apr 15st, All | Snapshot Apr 15th, Critical |
| --- | --- | --- | --- | --- | --- | --- |
| 1 | 3.79 | 2.07 | 3.52 | 1.83 | 3.78 | 2.73 |
| 2 | 4.09 | 2.03 | 2.03 | 1.79 | 1.73 | 1.97 |
| 3 | 5.99 | 1.81 | 4.20 | 1.19 | 3.37 | 3.72 |
| 4 | 6.35 | 1.58 | 5.46 | 0.81 | 3.39 | 2.06 |
| 5 | 5.32 | 1.41 | 2.19 | 1.22 | 4.89 | 1.17 |
| 6 | 3.48 | 2.14 | 2.32 | 2.27 | 2.84 | 2.11 |
| 7 | 3.94 | 1.04 | 2.32 | 0.71 | 1.63 | 1.03 |
| 8 | 4.86 | 1.33 | 3.14 | 1.90 | 3.39 | 1.04 |
| All | 4.11 | 1.29 | 2.41 | 0.75 | 2.68 | 1.25 |
| Mean (SE) | 4.72 (1.07) | 1.68 (0.40) | 3.15 (1.20) | 1.47 (0.56) | 3.13 (1.07) | 1.98 (0.93) |

**Table S10:** Random subset of patients holdout: ROC AUC and Brier Score of death prediction and critical-state-visit prediction by held-out group. Critical-state-visit prediction is not including patients started at critical state. The mean ROC AUC and Brier Scores estimates for death prediction over the eight held-out subsets are 0.955 (SE=0.035) and 0.043 (SE=0.011); the respective numbers of visiting-critical prediction are 0.880 (SE=0.040) and 0.049 (SE=0.013).

| group number | group size | Death Prediction |  |  | Visiting-Critical Prediction |  |  |
| --- | --- | --- | --- | --- | --- | --- | --- |
|  |  | number of deceased | AUC | Brier | number in critical (started in critical) | AUC | Brier |
| 1 | 330 | 26 | 0.966 | 0.047 | 20 (29) | 0.899 | 0.034 |
| 2 | 331 | 29 | 0.936 | 0.056 | 23 (28) | 0.862 | 0.048 |
| 3 | 329 | 19 | 0.980 | 0.038 | 32 (22) | 0.958 | 0.030 |
| 4 | 329 | 20 | 0.978 | 0.028 | 30 (23) | 0.860 | 0.051 |
| 5 | 331 | 24 | 0.977 | 0.030 | 34 (23) | 0.828 | 0.053 |
| 6 | 330 | 28 | 0.958 | 0.057 | 33 (17) | 0.904 | 0.072 |
| 7 | 330 | 19 | 0.876 | 0.038 | 22 (20) | 0.849 | 0.049 |
| 8 | 330 | 28 | 0.971 | 0.052 | 32 (22) | 0.876 | 0.055 |

**Table S11:** Hospital holdout: Mean absolute error (MAE) for each held-out hospital for predicting number of hospitalized and number of critical patients under the evaluation setups described in subsection S1.3.

| Hospital | Number of Patients | All Patients (MAE) | Critical Patients (MAE) | Snapshot Apr 1st All (MAE) | Snapshot Apr 1st Critical (MAE) | Snapshot Apr 15st All (MAE) | Snapshot Apr 15st Critical (MAE) |
| --- | --- | --- | --- | --- | --- | --- | --- |
| H1 | 298 | 8.51 | 2.24 | 3.57 | 1.44 | 4.57 | 1.19 |
| H2 | 166 | 6.48 | 0.82 | 4.10 | 0.66 | 3.83 | 1.15 |
| H3 | 142 | 3.91 | 1.55 | 1.57 | 0.64 | 1.14 | 1.29 |
| H4 | 105 | 3.78 | 1.01 | 1.65 | 1.96 | 1.53 | 1.46 |
| H5 | 343 | 5.55 | 1.67 | 2.68 | 1.29 | 5.74 | 2.08 |
| H6 | 111 | 2.11 | 0.75 | 2.02 | 0.63 | 2.15 | 0.92 |
| H7 | 373 | 11.44 | 7.06 | 7.11 | 8.58 | 10.40 | 7.74 |
| H8 | 215 | 5.71 | 2.61 | 7.79 | 1.10 | 2.41 | 3.66 |

**Table S12:**
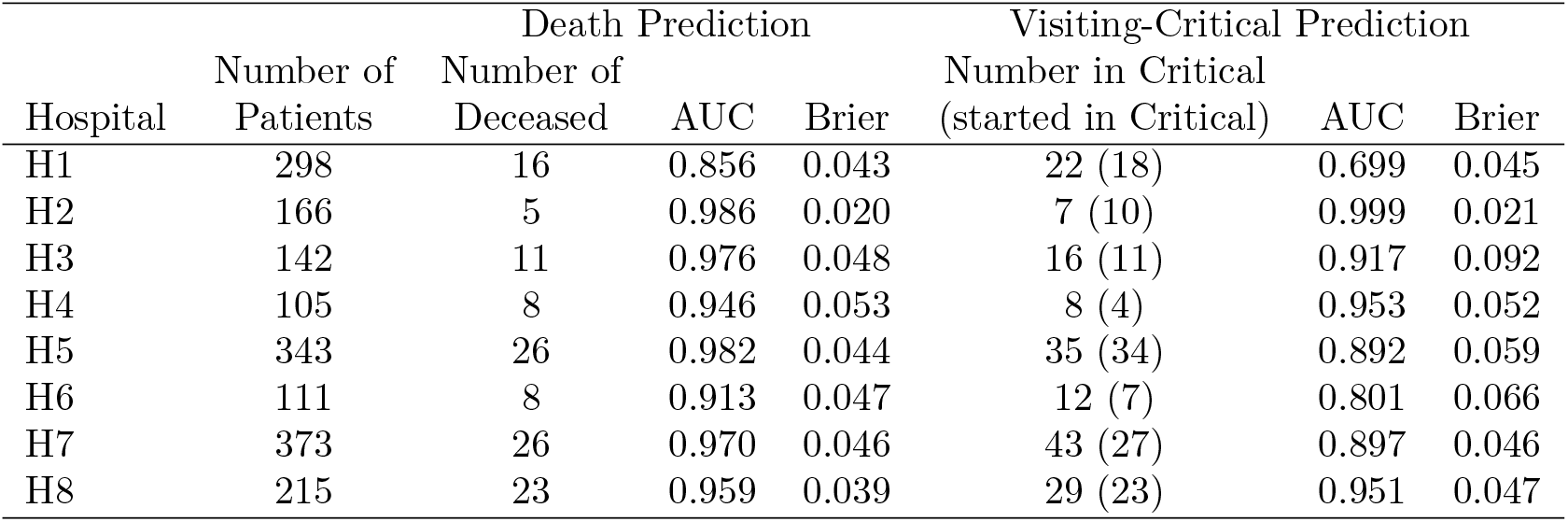
Hospital holdout: ROC AUC and Brier Score of death prediction and critical-state visit prediction for each held-out hospital. Critical-state-visit prediction does not include patients who started hospitalization at critical state.

**Table S13:** Predicted number of deaths (in-hospital mortality) within a random subset of 330 held-out patients: based on Arrival plus Snapshot prediction results of hypothetical scenarios, prediction starts on the 15th day of the observed arrival process. The numbers are the observed and predicted number of deaths for each hypothetical scenario from hospitalization day up to day *t* in hospital, *t* =5, 10*,…*, 35.

| Day $t$ | Observed | Expected | Younger | Milder | NH Outbreak |
| --- | --- | --- | --- | --- | --- |
| 5 | 7 | 6.5 | 0.6 | 3.7 | 8.2 |
| 10 | 16 | 16.6 | 1.9 | 11.7 | 22.6 |
| 15 | 20 | 20.4 | 2.5 | 15.0 | 28.3 |
| 20 | 23 | 22.8 | 3.0 | 17.3 | 32.1 |
| 25 | 24 | 25.4 | 3.4 | 19.7 | 36.5 |
| 30 | 25 | 25.9 | 3.5 | 20.1 | 37.0 |
| 35 | 26 | 26.6 | 3.7 | 20.6 | 37.7 |

**Figure S2:**
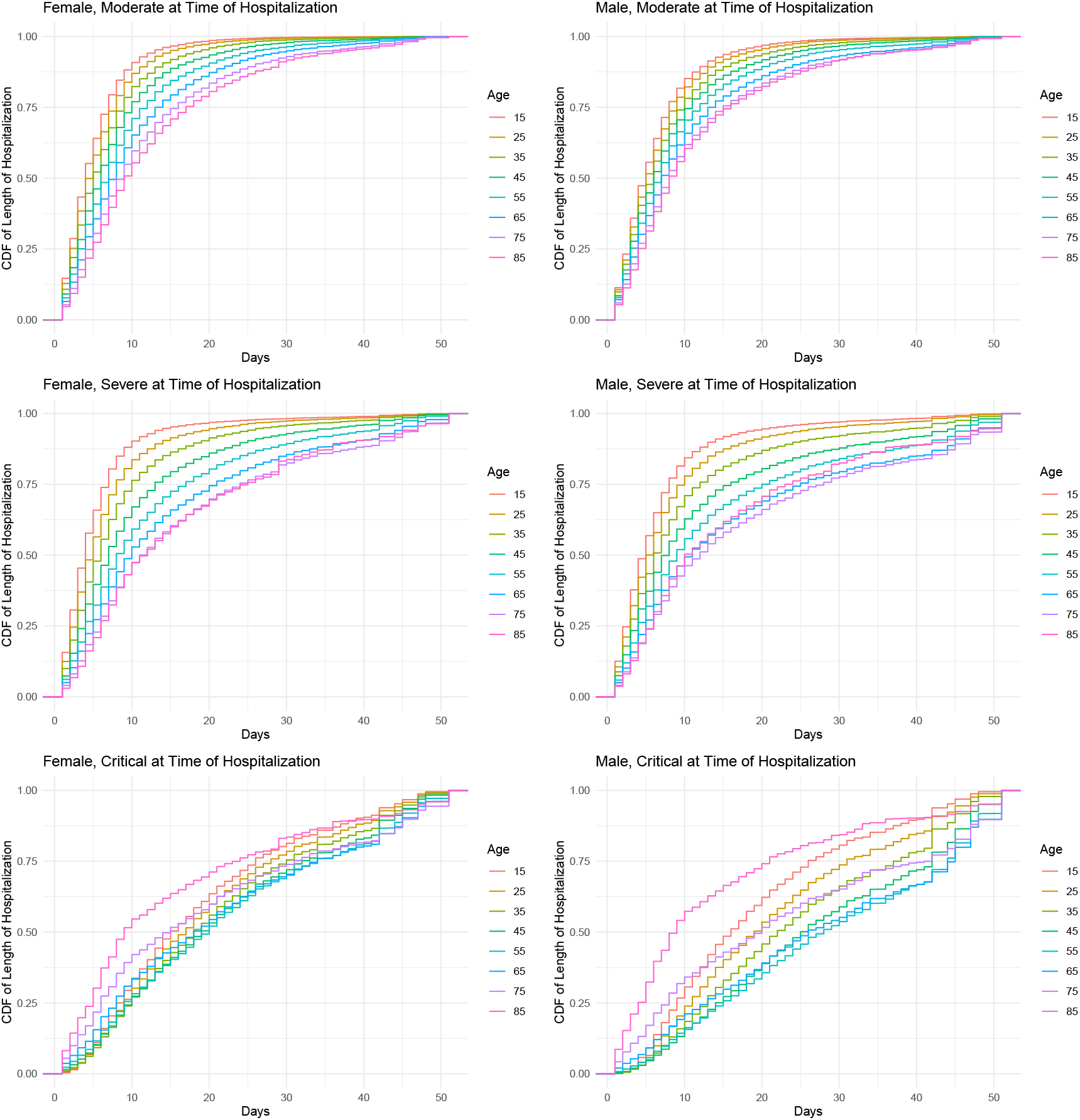
The cumulative distribution function of the length of hospitalization by patient types (sex, state at time of hospitalization and age). Each curve is based on 20,000 MC paths.

**Figure S3:**
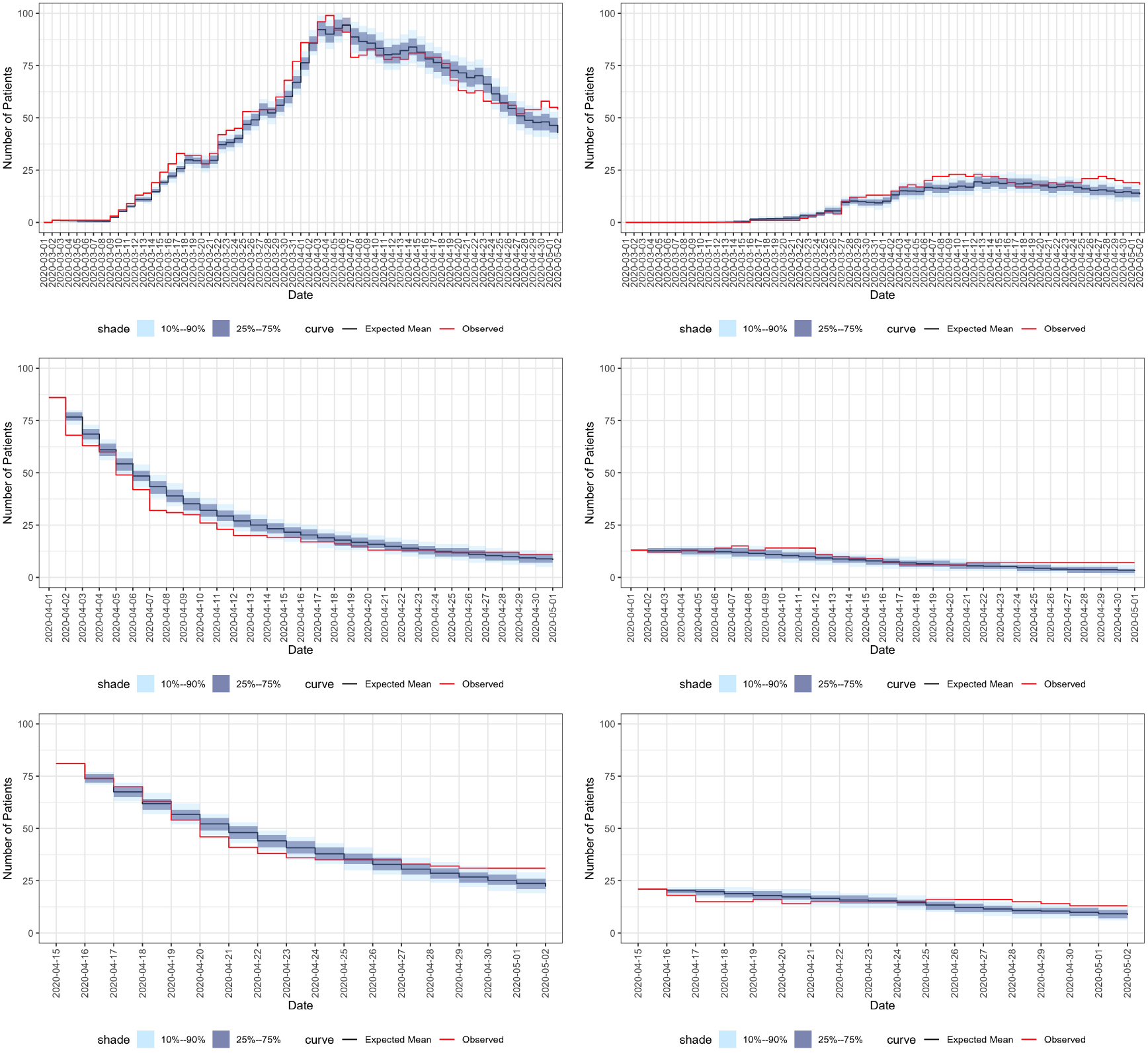
Prediction results of one held-out random group of patients. **Left figures**: utilization predictions for the entire held-out sample. **Right figures**: utilization predictions for critical patients among the held-out sample. **Top figures**: Arrival-type predictions of the entire held-out set based on the observed arrival process. **Middle figures**: Snapshot-type predictions for patients at the hospital on April 1st. **Bottom figures**: Snapshot predictions for patients in the hospital on April 15th. For description of Arrival and Snapshot see Section S1.3.

**Figure S4:**
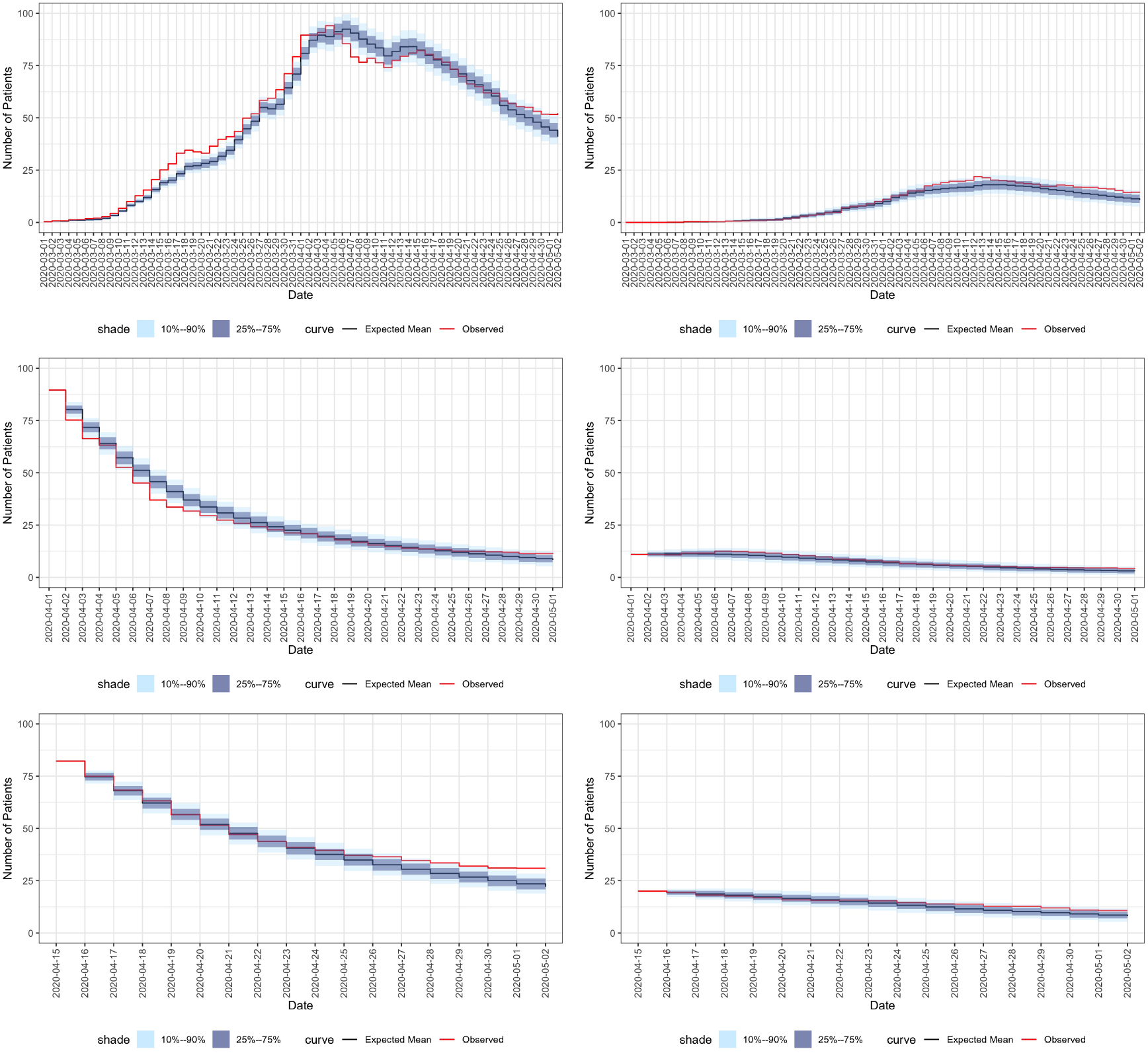
Prediction results summarized over all 8 held-out random groups of patients. **Left figures**: utilization predictions for the entire held-out sample. **Right figures**: utilization predictions for critical patients among the held-out sample. **Top figures**: Arrival-type predictions of the entire held-out set based on the observed arrival process. **Middle figures**: Snapshot-type predictions for patients at the hospital on April 1st. **Bottom figures**: Snapshot predictions for patients in the hospital on April 15th. For description of Arrival and Snapshot see Section S1.3.

**Figure S5:**
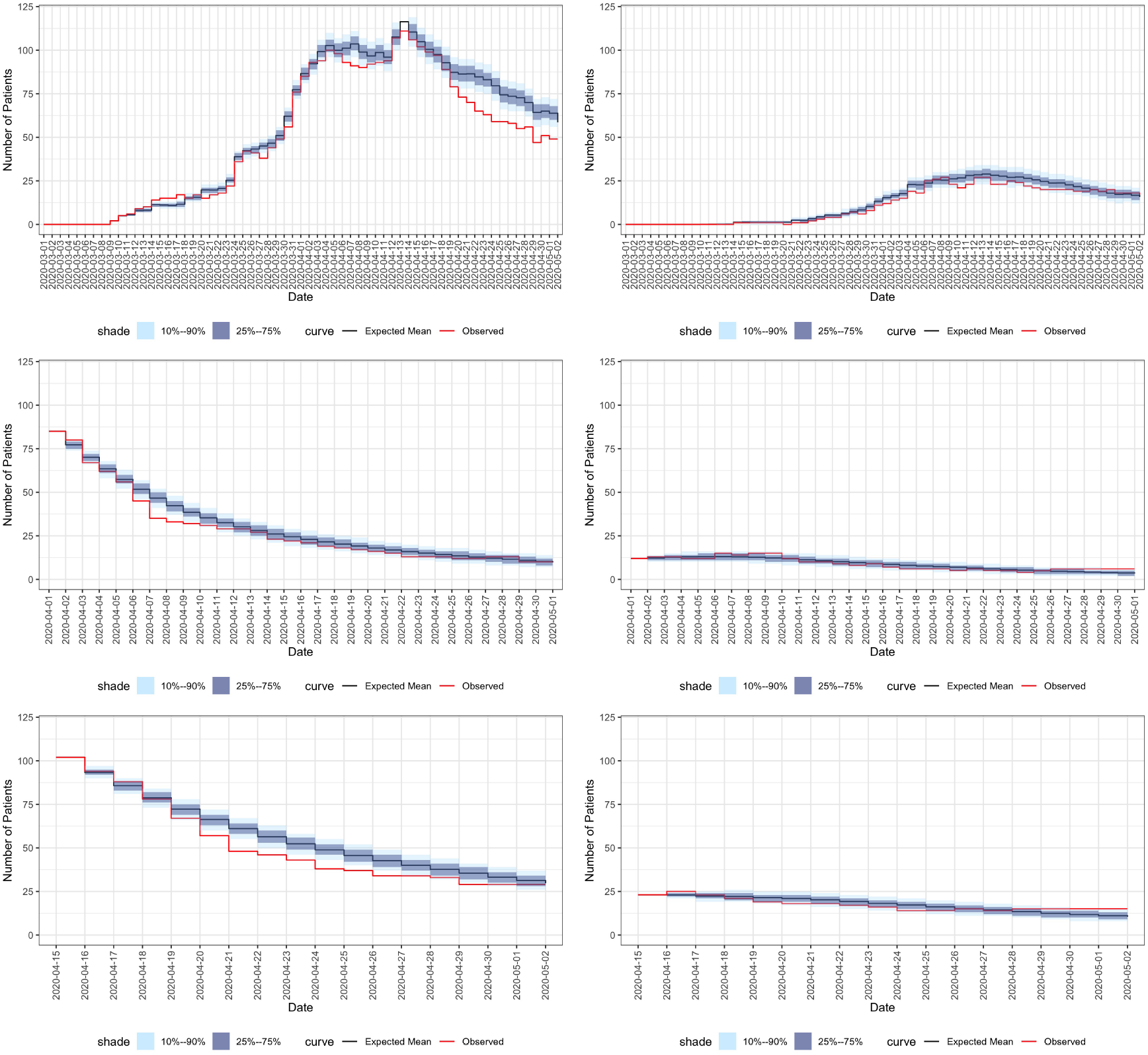
Prediction results -H5 Hospital is held out and predicted based on all other Israeli Hospitals. **Left figures**: utilization predictions for the entire held-out sample. **Right figures**: utilization predictions for critical patients among the held-out sample. **Top figures**: Arrival-type predictions of the entire held-out set based on the observed arrival process. **Middle figures**: Snapshot-type predictions for patients at the hospital on April 1st. **Bottom figures**: Snapshot predictions for patients in the hospital on April 15th. For description of Arrival and Snapshot see S1.3.

**Figure S6:**
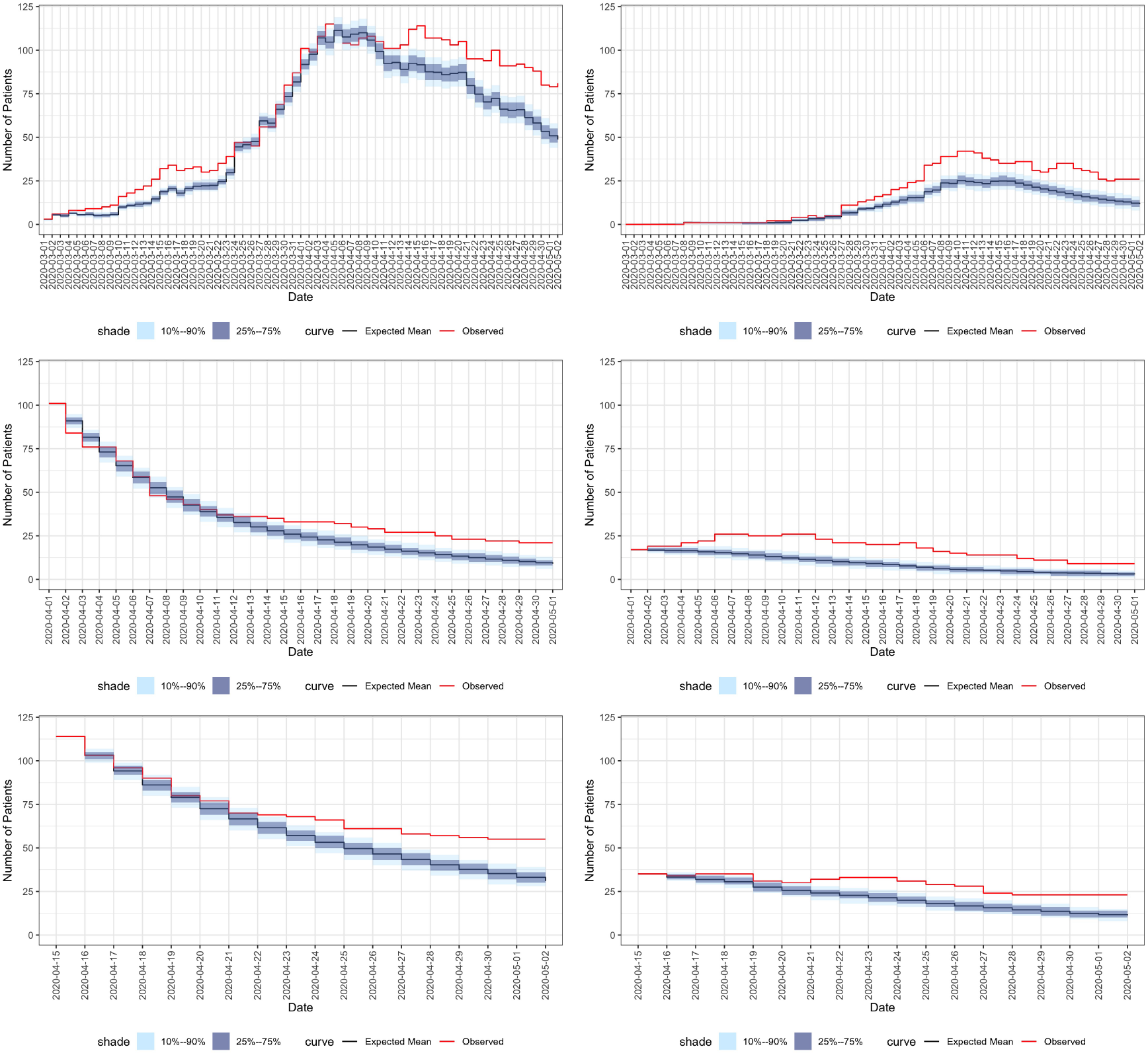
Prediction results -H7 Hospital is held out and predicted based on all other Israeli Hospitals. **Left figures**: utilization predictions for the entire held-out sample. **Right figures**: utilization predictions for critical patients among the held-out sample. **Top figures**: Arrival-type predictions of the entire held-out set based on the observed arrival process. **Middle figures**: Snapshot-type predictions for patients at the hospital on April 1st. **Bottom figures**: Snapshot predictions for patients in the hospital on April 15th. For description of Arrival and Snapshot see S1.3.

**Figure S7:**
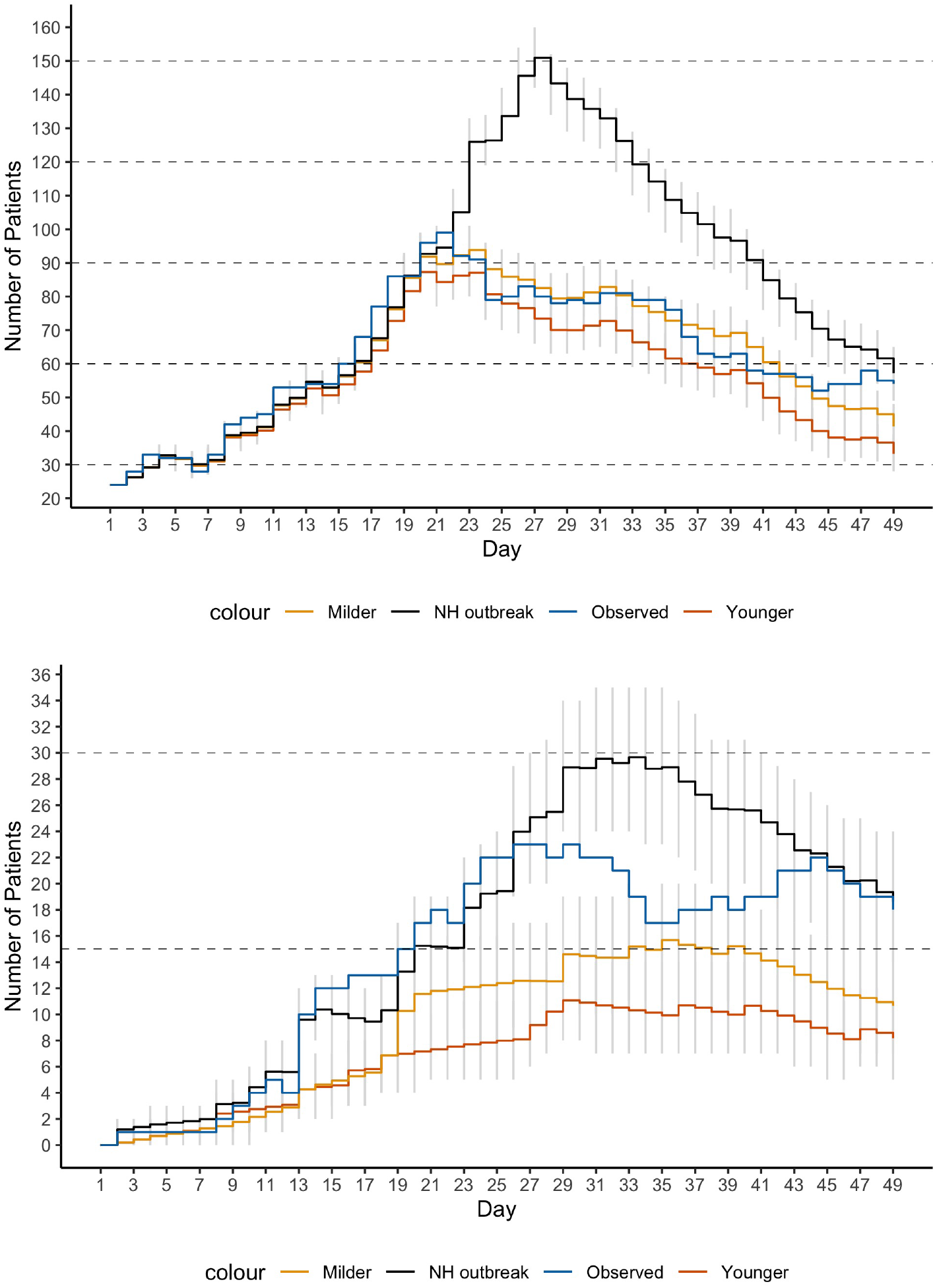
Arrival plus Snapshot prediction results of hypothetical scenarios: 330 random patients were held-out and prediction started on the 15th day of the observed arrival process (denoted as Day 1 in the figures). **Top figure**: utilization predictions based on the entire held-out sample. **Bottom figure**: utilization predictions for critical patients among the held-out sample. Gray vertical lines are point-wise 10%-90% confidence predictions.

## Footnotes

a https://github.com/JonathanSomer/covid-19-multi-state-model

b https://covid19-hospitalcourse.net/

c In addition, 30 patients were either dead on arrival or deceased within their first hospitalization day, and five were discharged on their first hospitalization day.

